# Time trends in new diagnoses of 19 long-term conditions: a population-level cohort study in England using OpenSAFELY

**DOI:** 10.1101/2025.07.11.25331090

**Authors:** Mark D Russell, Andrea Schaffer, Katie Bechman, Mark Gibson, Jon Massey, Rose Higgins, Brian MacKenna, Peter Inglesby, Seb Bacon, Amir Mehrkar, Ben Goldacre, Edward Alveyn, Victoria Allen, Zijing Yang, Samir Patel, Maryam A Adas, Gurjinder Sandhu, Elizabeth Price, Rouvick M Gama, Kate Bramham, Matthew Hotopf, Sam Norton, Andrew P Cope, James B Galloway

## Abstract

**Objectives:** To evaluate temporal changes in the incidence and prevalence of 19 long-term conditions in England, quantifying the impact of the COVID-19 pandemic on diagnosis rates by disease, age group, sex, socioeconomic status, and ethnicity.

**Design:** Observational cohort study.

**Setting:** Primary care and hospital admission data, with the approval of NHS England.

**Participants:** 27,132,190 individuals registered with general practices in England contributing data to the OpenSAFELY-TPP platform.

**Main outcomes measures:** Temporal trends in age and sex-standardised incidence and prevalence were evaluated for 19 long-term conditions between April 1, 2016, and November 30, 2024. Differences between expected and observed incidence rates after the onset of the COVID-19 pandemic were compared using seasonal autoregressive integrated moving-average models.

**Results:** Between March 2020 and November 2024, persistent large deficits in incident diagnoses were evident for depression (738,068 [28.0%] fewer diagnoses than expected; 95% CI 701,452 to 774,685), asthma (150,708 [16.0%] fewer diagnoses; 95% CI 133,300 to 168,117), COPD (84,084 [15.1%] fewer diagnoses; 95% CI 74,342 to 93,827), osteoporosis (78,891 [16.5%] fewer diagnoses; 95% CI 72,804 to 84,978) and psoriasis (56,231 [17.6%] fewer diagnoses; 95% CI 51,054 to 61,407). Conversely, post-pandemic diagnoses of chronic kidney disease (CKD) have increased by 32.7% above expected levels, corresponding to 325,996 additional diagnoses (95% CI 252,212 to 399,779). Dementia diagnoses have rebounded above pre-pandemic levels for individuals of White ethnicity and less deprived socioeconomic quintiles, but remain lower than expected for individuals from other ethnicities and more deprived communities.

**Conclusions:** There has been a lasting and disproportionate impact of the pandemic on conditions including depression, asthma, COPD and osteoporosis, contrasting a post-pandemic surge in CKD diagnoses. Analyses stratified by ethnicity and socioeconomic status reveal inequity in the recovery from the pandemic, particularly for individuals with dementia. Importantly, this study demonstrates the potential for near real-time monitoring of disease epidemiology using routinely collected health data, informing strategies to enhance case detection and address healthcare disparities.

**Summary box:** *What is already known:*

- Early in the COVID-19 pandemic, large deficits in newly-recorded diagnoses were reported for a range of long-term conditions, including chronic obstructive pulmonary disease (COPD), depression, asthma and rheumatoid arthritis.
- However, no population-level studies have evaluated whether diagnosis rates have subsequently recovered as healthcare services emerge from the pandemic, or compared the impact by disease, age group, sex, socioeconomic status, and ethnicity.

*What this study adds:*

- Large deficits in diagnoses remain evident for conditions including depression, COPD, asthma, osteoporosis and psoriasis, contrasting a surge in new diagnoses of chronic kidney disease since 2022.
- Diagnosis rates for dementia have reached pre-pandemic levels for individuals of White ethnicity and those from less deprived socioeconomic communities, but remain lower than expected among individuals from other ethnicities and more deprived communities.
- This study demonstrates how routinely collected health data can transform monitoring of disease epidemiology in near real-time and highlight healthcare inequity.

## Introduction

The COVID-19 pandemic had an unprecedented impact on healthcare systems worldwide, disrupting service delivery through redeployment of staff, implementation of stay-at-home guidance, and abrupt shifts to healthcare utilisation.^1-4^ The consequences of these disruptions on care for long-term health conditions remains poorly understood.

The OpenSAFELY platform enables primary care and hospital data to be analysed in a secure environment for up to 58 million people, covering 99% of England’s population.^5^ In proof-of-concept studies, data in OpenSAFELY were used to highlight marked decreases in new autoimmune inflammatory arthritis diagnoses (e.g. rheumatoid arthritis) and gout during the early pandemic.^6,7^ While incidence rates for these conditions returned to pre-pandemic levels during the second year of the pandemic, no compensatory increase in diagnoses above pre-pandemic levels had been observed as of 2023, indicating that a substantial proportion of cases remained undiagnosed at that point.^6-8^

It remains unclear whether there has been a subsequent recovery in diagnoses as we emerge from the pandemic, or to what extent other long-term conditions have been affected. Our objective was to evaluate changes in the recorded incidence and prevalence of 19 long-term conditions, spanning multiple disease areas, using population-level data in OpenSAFELY. We sought to quantify the relative and absolute impact of the pandemic on diagnosis rates, stratified by disease, age group, sex, socioeconomic status, and ethnicity.

## Methods

### Study design and data source

A population-level, observational cohort study was performed using linked primary and secondary care data in the OpenSAFELY-TPP platform, which contains health data for individuals registered with general practices in England using TPP SystmOne software.^5^ OpenSAFELY-TPP is representative of England’s population in terms of age, sex, socioeconomic status, ethnicity, and causes of death.^9^ Primary care records were linked to the Hospital Episode Statistics dataset through OpenSAFELY.

### Study population and case definitions

The study period was from April 1, 2016, to November 30, 2024. The reference population consisted of all individuals registered with TPP practices in England at any point during the study period who did not have type 1 opt-outs registered with their practice (type 1 opt-outs indicate individuals who do not want their personal data being shared with any organisation not involved in their direct care, representing approximately 3% of the population of England in 2017^10,11^).

Incidence and prevalence were reported for the following conditions during the study period: asthma, chronic obstructive pulmonary disease (COPD), coronary heart disease, stroke or transient ischaemic attack (TIA), type 2 diabetes mellitus, chronic kidney disease stages 3 to 5 (CKD), ulcerative colitis, Crohn’s disease, coeliac disease, dementia, multiple sclerosis, epilepsy, depression, psoriasis, atopic dermatitis, osteoporosis, polymyalgia rheumatica, and rheumatoid arthritis. These conditions were chosen as examples of long-term health conditions spanning a range of disease areas, to enable comparisons of the relative impact of the pandemic on different diseases. Incident cases were defined as the first appearance of a diagnostic code for each condition in individuals’ primary care or hospital admission records (see Appendix for codelists). Hospitalisations where relevant diagnostic codes were listed as the primary cause for that admission were included in estimates of incidence or prevalence; however, hospitalisations where diagnostic codes were listed in secondary positions were not included, due to less reliable coding. For example, the inclusion of secondary admission codes for rheumatoid arthritis resulted in incidence rates that were 3-fold higher than estimates from previous population-level studies.^6,12^ In our study, at least 12 months of continuous registration prior to diagnosis was required to ensure only incident diagnoses were captured.

Monthly incidence rates were calculated for each condition by dividing the number of individuals with index diagnoses in each calendar month, by the number of individuals in the reference population without previously recorded diagnostic codes for that condition who had at least 12 months of continuous registration at the start of each month. Direct age and sex standardisation was performed with reference to the 2013 European Standard Population,^13^ using 10-year age bands up to the age of 80 years. Ten-year age bands were selected over 5-year age bands due to the computationally-intensive nature of deriving monthly numerator and denominator data for more than 27 million individuals. For comparison, age and sex-standardised incidence rates were presented alongside rates without standardisation. Monthly incidence rates were presented per 100,000 population alongside 3-monthly rolling averages, representing the mean of the current, previous and subsequent months. Incidence rates for each condition were reported separately by subgroups of sex, age band, socioeconomic status (defined as quintiles of Index of Multiple Deprivation [IMD]^14^), and ethnicity (categorised into White, Mixed, Asian or Asian British, Black or Black British, or Chinese or other ethnic groups^15^). No age or sex-standardisation was performed for subgroup analyses by IMD or ethnicity for computational reasons.

Temporal trends in the annual prevalence of each condition were estimated by dividing the number of people with prevalent diagnostic codes on April 1 of each year of the study period, by the number of individuals currently registered at the same time-point. Age and sex-standardised estimates of prevalence per 100,000 population were presented in addition to estimates without standardisation.

To explore potential reasons for the observed decline in depression diagnoses during the study period, a sensitivity analysis was performed using an expanded depression codelist, which included diagnostic codes indicative of depressive symptoms (e.g. depressed mood), as well as confirmed depression diagnoses (see Appendix).

### Statistical methods

Baseline sociodemographic characteristics were tabulated without inferential statistics for the reference population and, separately, for individuals with incident diagnoses of each condition during the study period.

Differences between expected and observed incidence rates for each condition after the onset of the COVID-19 pandemic were estimated using Seasonal Autoregressive Integrated Moving Average (SARIMA) models – a forecasting method that accounts for underlying trends and seasonal patterns in time-series data (see Appendix for further methodological details).^16,17^ Monthly incidence data from April, 2016, to February, 2020 (i.e. prior to the onset of the first COVID-19 lockdown in England) were fitted and used to predict expected incidence rates for the period March, 2020, to November, 2024. Absolute and relative differences between expected and observed incidence rates were calculated and presented for the full post-pandemic period (March, 2020, to November, 2024) and separately for the following periods: March, 2020, to February, 2021; March, 2021, to February, 2022; March, 2022, to February, 2023; and March, 2023, to November, 2024. Absolute differences in numbers of recorded diagnoses with reference to the full population of England were estimated by extrapolating differences in expected vs. observed incidence rates for the study population to mid-year population estimates for England.^18^

Python 3.11 was used for data management; Stata version 18 and R version 4.41 were used for statistical analyses. Code for data management and analysis, as well as codelists, are archived online (https://github.com/opensafely/disease_incidence). As analyses were primarily descriptive in nature, no correction for multiple hypothesis testing was performed. For statistical disclosure control, frequency counts were rounded to the nearest 5 and non-zero counts below 8 were redacted.

### Study approval and ethics

Approval to undertake this study under the remit of service evaluation was obtained from King’s College Hospital NHS Foundation Trust, London. No further ethical approval was required as per UK Health Research Authority guidance.

### Patient and public involvement

OpenSAFELY has involved patients and the public in various ways, including: a public website that provides a detailed description of the platform in language suitable for a lay audience (https://opensafely.org); participation in two citizen juries exploring public trust in OpenSAFELY; co-development of an explainer video (https://www.opensafely.org/about/); patient representation, who are experts by experience, on our OpenSAFELY Oversight Board; partnering with Understanding Patient Data to produce lay explainers on the importance of large datasets for research; presentations at various online public engagement events to key communities (e.g. Healthcare Excellence Through Technology; Faculty of Clinical Informatics annual conference; NHS Assembly; HDRUK symposium); and more. To ensure the patient voice is represented, OpenSAFELY are working closely to decide on language choices with appropriate medical research charities (e.g. Association of Medical Research Charities). Information and interpretation of our findings will be shared through press releases, social media channels, and plain language summaries.

## Results

Between April 1, 2016, and November 30, 2024, 27,132,190 individuals in England had data available for analysis in OpenSAFELY-TPP. Sociodemographic data for these individuals, and for individuals who developed incident diagnoses of 19 long-term conditions, are shown in Supplementary Table S1.

Relative to pre-pandemic trends, incidence rates for all 19 conditions decreased sharply after the onset of the COVID-19 pandemic; however, the magnitude of these decreases varied substantially by condition (Supplementary Figures S1 and S2). Observed incidence rates after March 2020 were compared to expected incidence rates (modelled using SARIMA), to quantify relative and absolute deficits in diagnoses (Figure 1 and Table 1; Supplementary Table S2). During the first year of the pandemic, the largest relative deficits in incidence rates were evident for COPD (−55.6%; 95% CI (−56.9 to −54.1), psoriasis (−43.8%; 95% CI (−45.5 to −41.9), atopic dermatitis (−39.1%; 95% CI −43.4 to −34.0), osteoporosis (−36.8%; 95% CI −38.6 to −35.0), coeliac disease (−32.0%; 95% CI −35.2 to −28.4) and asthma (−31.8%; 95% CI −34.0 to −29.3). The smallest relative deficits in incidence rates were observed for stroke and TIA (−7.06%; 95% CI −8.75 to −5.31), and polymyalgia rheumatica (−8.09%; 95% CI −12.1 to −3.73).

**Table 1.** Differences between expected and observed incidence rates for 19 long-term conditions after the onset of the COVID-19 pandemic in England. Expected and observed incidence rates for each condition are compared during the first year after the onset of the COVID-19 pandemic (March 1, 2020, to February 28, 2021) and during the full post-pandemic study period (March 1, 2020, to November 30, 2024). Expected incidence rates were modelled using seasonal autoregressive integrated moving averages (SARIMA), utilising data from April, 2016, to February, 2020. The relative differences between the number of expected vs. observed diagnoses are shown, in addition to the absolute differences in expected vs. observed diagnoses when extrapolated to the full population of England. COPD: chronic obstructive pulmonary disease; TIA: transient ischaemic attack.

| Condition | First year of pandemic (March 2020 to February 2021) |  |  | Full post-pandemic period (March 2020 to November 2024) |  |  |  |
| --- | --- | --- | --- | --- | --- | --- | --- |
|  | Observed incidence per 100,000 population | Expected incidence per 100,000 population (95% CI) | Relative difference between expected and observed (95% CI) | Observed incidence per 100,000 population | Expected incidence per 100,000 population (95% CI) | Relative difference between expected and observed (95% CI) | Absolute difference in diagnoses for the full population of England (95% CI) |
| Asthma | 238.1 | 348.9<br>(336.9, 360.9) | -31.8%<br>(-34.0, -29.3) | 1,388 | 1,652<br>(1,622, 1,683) | -16.0%<br>(-17.5, -14.4) | -150,708<br>(-168,117, -133,300) |
| Atopic Dermatitis | 41.2 | 67.5<br>(62.4, 72.7) | -39.1%<br>(-43.4, -34.0) | 269.1 | 261.3<br>(231.2, 291.4) | 2.99%<br>(-7.64, 16.4) | 4,460<br>(-12,705, 21,625) |
| Coronary Heart Disease | 234.3 | 287.0<br>(281.1, 292.9) | -18.4%<br>(-20.0, -16.6) | 1249 | 1,339<br>(1,322, 1,356) | -6.71%<br>(-7.87, -5.52) | -51,251<br>(-60,843, -41,660) |
| Chronic Kidney Disease | 271.6 | 363.9<br>(340.0, 387.9) | -25.4%<br>(-30.0, -20.1) | 2,322 | 1,750<br>(1,621, 1,879) | 32.7%<br>(23.5, 43.2) | 325,996<br>(252,212, 399,779) |
| Coeliac Disease | 17.8 | 26.1<br>(24.8, 27.4) | -32.0%<br>(-35.2, -28.4) | 119.7 | 123.8<br>(120.8, 126.8) | -3.31%<br>(-5.58, -0.94) | -2,340<br>(-4,033, -648) |
| COPD | 96.8 | 218.0<br>(211.1, 224.8) | -55.6%<br>(-56.9, -54.1) | 825.7 | 973.1<br>(956.0, 990.2) | -15.1%<br>(-16.6, -13.6) | -84,084<br>(-93,827, -74,342) |
| Crohn's Disease | 15.7 | 19.0<br>(18.0, 20.0) | -17.7%<br>(-21.8, -13.2) | 88.4 | 88.3<br>(84.1, 92.6) | 0.037%<br>(-4.55, 5.09) | 18.8<br>(-2,404, 2,441) |
| Dementia | 150.1 | 194.5<br>(188.3, 200.6) | -22.8%<br>(-25.2, -20.3) | 887.2 | 924.9<br>(909.2, 940.7) | -4.08%<br>(-5.68, -2.42) | -21,516<br>(-30,479, -12,553) |
| Depression | 651.0 | 924.7<br>(896.7, 952.7) | -29.6%<br>(-31.7, -27.4) | 3,329 | 4,623<br>(4,558, 4,687) | -28.0%<br>(-29.0, -27.0) | -738,068<br>(-774,685, -701,452) |
| Diabetes Mellitus Type 2 | 361.4 | 462.1<br>(434.0, 490.2) | -21.8%<br>(-26.3, -16.7) | 2,344 | 2,192<br>(2,107, 2,278) | 6.90%<br>(2.90, 11.2) | 86,274<br>(37,686, 134,863) |
| Epilepsy | 38.6 | 43.7<br>(42.0, 45.4) | -11.7%<br>(-15.1, -8.08) | 193.9 | 207.0<br>(202.1, 211.8) | -6.30%<br>(-8.44, -4.05) | -7,435<br>(-10,199, -4,671) |
| Heart Failure | 204.6 | 252.7<br>(246.2, 259.2) | -19.0%<br>(-21.0, -16.9) | 1,203 | 1,250<br>(1,231, 1,268) | -3.71%<br>(-5.13, -2.25) | -26,461<br>(-37,102, -15,821) |
| Multiple Sclerosis | 6.90 | 7.78<br>(7.35, 8.22) | -11.4%<br>(-16.1, -6.21) | 34.4 | 36.7<br>(35.5, 37.9) | -6.24%<br>(-9.18, -3.11) | -1,305<br>(-1,982, -629) |
| Osteoporosis | 108.6 | 172.0<br>(167.2, 176.8) | -36.8%<br>(-38.6, -35.0) | 698.3 | 836.6<br>(825.9, 847.3) | -16.5%<br>(-17.6, -15.5) | -78,891<br>(-84,978, -72,804) |
| Polymyalgia Rheumatica | 45.1 | 49.1<br>(46.8, 51.3) | -8.09%<br>(-12.1, -3.73) | 209.6 | 220.7<br>(213.7, 227.6) | -5.0%<br>(-7.91, -1.91) | -6,298<br>(-10,267, -2,329) |
| Psoriasis | 69.1 | 122.8<br>(118.9, 126.8) | -43.8%<br>(-45.5, -41.9) | 461.9 | 560.5<br>(551.4, 569.5) | -17.6%<br>(-18.9, -16.2) | -56,231<br>(-61,407, -51,054) |
| Rheumatoid Arthritis | 30.9 | 40.9<br>(39.1, 42.8) | -24.5%<br>(-27.7, -21.0) | 174.8 | 194.7<br>(190.5, 198.8) | -10.2%<br>(-12.1, -8.26) | -11,344<br>(-13,717, -8,971) |
| Stroke/TIA | 230.7 | 248.2<br>(243.7, 252.8) | -7.06%<br>(-8.75, -5.31) | 1,178 | 1,158<br>(1,146, 1,171) | 1.68%<br>(0.58, 2.80) | 11,101<br>(3,901, 18,301) |
| Ulcerative Colitis | 25.7 | 29.6<br>(28.3, 31.0) | -13.4%<br>(-17.1, -9.26) | 143.4 | 140.7<br>(137.6, 143.9) | 1.92%<br>(-0.32, 4.26) | 1,538<br>(-264, 3,340) |

**Figure 1.**
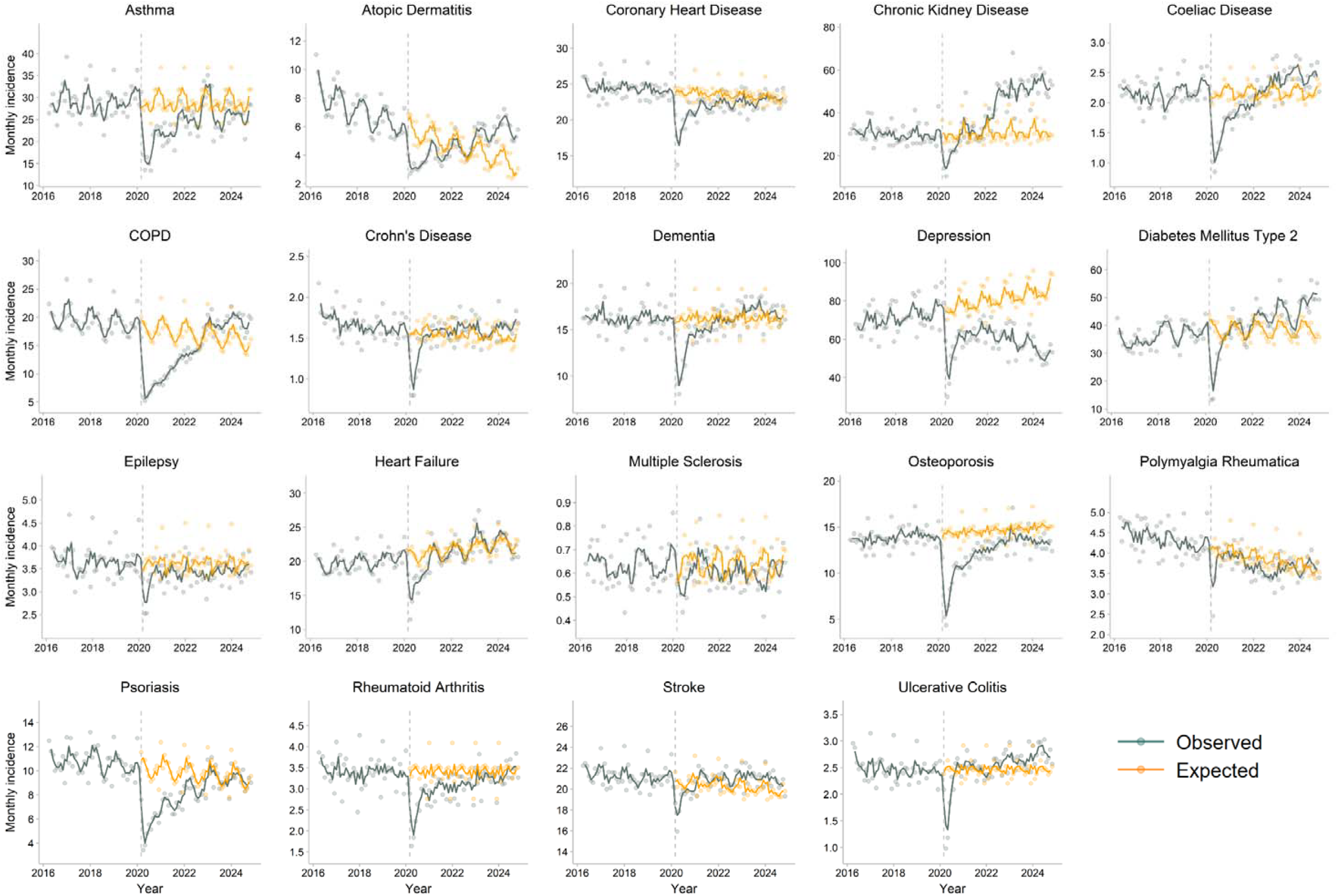
Comparison between observed (dark green) and expected (orange) incidence rates for 19 long-term conditions after the onset of the COVID-19 pandemic in England. Expected incidence rates after March 2020 (vertical dashed line) were estimated using seasonal autoregressive integrated moving averages (SARIMA) models, utilising data from April 1, 2016, to February 28, 2020. Individual data points, representing age and sex-adjusted monthly incidence rates per 100,000 population, are presented alongside 3-monthly rolling averages of the current, preceding and subsequent months.

By November 2024, persistent deficits in incident diagnoses remained evident for several conditions (Table 1). Depression had the largest overall deficit in diagnoses between March 2020 and November 2024 (−28.0%; 95% CI −29.0 to −27.0). After a 29.6% decrease in new diagnoses during the first year of the pandemic (95% CI 27.4 to 31.7), the incidence of depression partly recovered, but remained below pre-pandemic levels, and was followed by a further decline in incidence between 2022 and 2024 (Figure 1). When extrapolated to the full population of England, there were 738,068 [28.0%] fewer diagnoses of depression than expected between March 2020 and November 2024 (95% CI 701,452 to 774,685). To explore potential reasons for the observed decline in depression diagnoses during the study period, a sensitivity analysis was performed using an expanded depression codelist, which included diagnostic codes indicative of depressive symptoms (e.g. depressed mood), as well as confirmed depression diagnoses (Supplementary Figure S3). The relative post-pandemic deficit in depression diagnoses was attenuated when these additional codes were included (−21.1%; 95% CI −22.2 to −19.9); however, the absolute deficit in diagnoses was comparable (740,479 fewer diagnoses; 95% CI 689,062 to 791,896) – reflective of the higher incidence rate for depression when including these additional codes.

The next largest absolute deficits in diagnoses were evident for asthma (150,708 [16.0%] decrease; 95% CI 133,300 to 168,117), COPD (84,084 [15.1%] decrease; 95% CI 74,342 to 93,827), osteoporosis (78,891 [16.5%] decrease; 95% CI 72,804 to 84,978), psoriasis (56,231 [17.6%] decrease; 95% CI 51,054 to 61,407) and coronary heart disease (51,251 [6.71%] decrease; 95% CI 41,660 to 60,843) (Table 1). In contrast, diagnosis rates for other conditions rebounded above pre-pandemic levels after the initial decreases – most notably for CKD (Table 1). Following a 25.4% decrease (95% CI 20.1 to 30.0) in new diagnoses during the first year of the pandemic, the incidence of CKD recovered to pre-pandemic levels by late 2020, before surging above pre-pandemic levels from 2022 onwards (Figure 1). Between March 2020 and November 2024, the recorded incidence of CKD was 32.7% higher than expected, based upon pre-pandemic trends (95% CI 23.5 to 43.2); corresponding to 325,996 additional CKD diagnoses in England (95% CI 252,212 to 399,779).

When analysed separately by sex (Figure 2), age group (Supplementary Figure S4), socioeconomic status (Figure 3), and ethnicity (Supplementary Figure S5), several disparities were evident in the post-pandemic recovery of incidence rates. For CKD, much of the post-pandemic increase in diagnoses occurred in individuals aged 80 years and over, and in those from less deprived socioeconomic quintiles, with comparable relative increases observed for individuals of White, Black and Asian ethnicities. For depression, the post-pandemic decrease in diagnoses occurred predominantly in individuals aged 20 to 39 years, in those of White or Mixed ethnicity, with comparable decreases across socioeconomic quintiles. For dementia, rebound increases in diagnoses above pre-pandemic levels were evident for individuals of White ethnicity and those from less deprived socioeconomic quintiles, but not for individuals of other ethnicities or those from more deprived socioeconomic quintiles.

**Figure 2.**
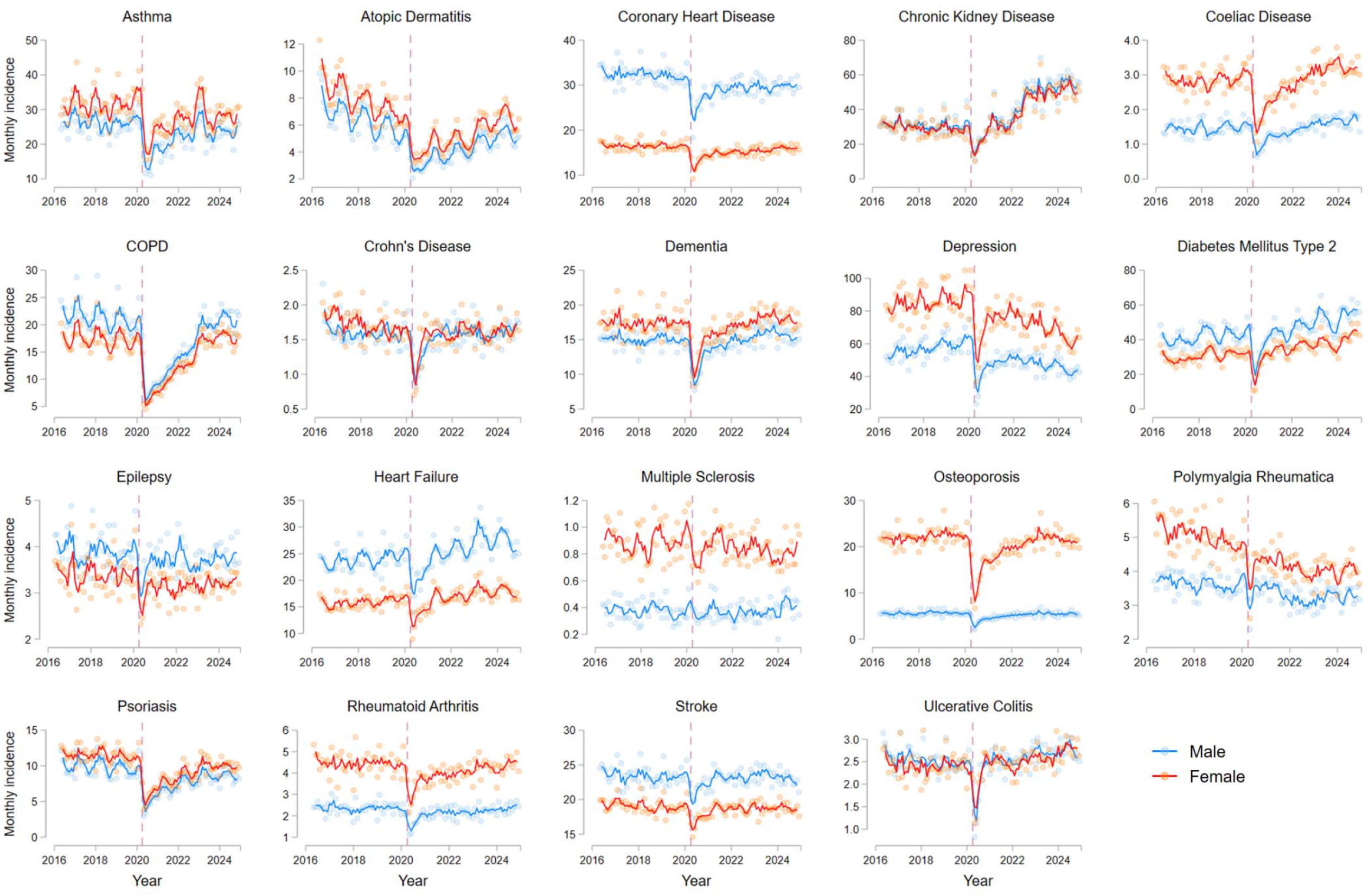
Monthly incidence rates for 19 long-term conditions in England between April 1, 2016, to November 30, 2024. Age-adjusted incidence rates are shown per 100,000 population, separated by sex. Individual data points, representing age-adjusted incidence rates for each month, are presented alongside 3-monthly rolling averages of the current, preceding and subsequent months. The vertical dashed line corresponds to the onset of the first COVID-19 lockdown in England (March 2020).

**Figure 3.**
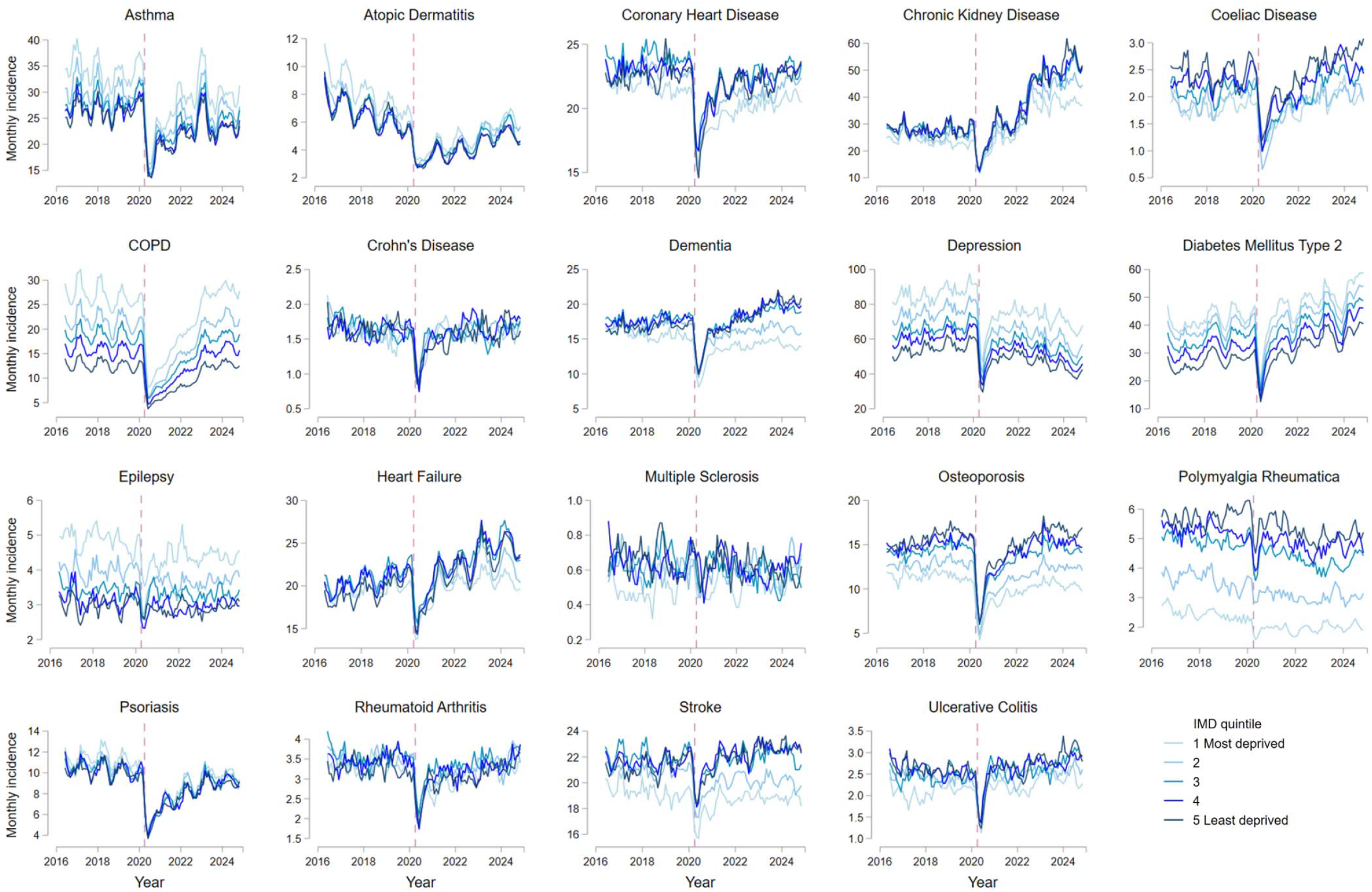
Monthly incidence rates for 19 long-term conditions in England between April 1, 2016, to November 30, 2024, separated by socioeconomic status. Socioeconomic status was divided into quintiles of Index of Multiple Deprivation (IMD), from 1 (most deprived quintile) to 5 (least deprived quintile). Incidence rates per 100,000 population are presented as 3-monthly rolling averages of the current, preceding and subsequent months. No age or sex-standardisation was performed due to computational requirements. The vertical dashed line corresponds to the onset of the first COVID-19 lockdown in England (March 2020).

Notable changes in the prevalence of several conditions were evident after the onset of the pandemic when compared with pre-pandemic trends (Supplementary Figures S6 and S7). A gradually declining pre-pandemic prevalence of dementia accelerated during the first year of the pandemic, before stabilising from 2021 onwards. Having been stable prior to the pandemic, the prevalence of COPD decreased after 2020, corresponding to the observed fall in incidence. In contrast, the declining pre-pandemic prevalence of CKD reversed abruptly after 2022, corresponding to the marked post-pandemic increase in new diagnoses.

## Discussion

Utilising data for 27 million individuals in England, our study highlights marked variation in the impact of the COVID-19 pandemic on diagnosis rates across a range of diseases and sociodemographic characteristics. New diagnoses of all 19 long-term conditions fell sharply during the first year of the pandemic, but with a disproportionate impact on conditions including COPD, asthma, psoriasis and osteoporosis. Rebound increases in diagnoses were subsequently observed for several conditions – particularly CKD, for which reported cases have surged since 2022. In contrast, substantial deficits in diagnoses have persisted for other conditions nearly five years after the onset of the pandemic, most notably for depression. Analyses stratified by ethnicity and socioeconomic status highlighted compensatory increases in dementia diagnoses for individuals of White ethnicity and those from less deprived socioeconomic quintiles, but not for individuals of other ethnicities or more deprived socioeconomic backgrounds. Importantly, this study demonstrates how routinely collected health data can transform how we monitor disease epidemiology in close to real-time.

Data from early in the pandemic reported sharp decreases in incident diagnoses for several long-term conditions, including asthma, COPD and rheumatoid arthritis.^6,7,17,19,20^ Little was known, however, about the extent of recovery in diagnosis rates as healthcare services emerged from the pandemic. Demonstrating this at a population level requires regularly updated data from primary and secondary care settings, which has only recently become possible with the introduction of the OpenSAFELY Trusted Research Environment in England.^5^ Our demonstration of a disproportionate impact of the early pandemic on respiratory conditions, such as COPD and asthma, supports the findings from a previous population-level study in Wales, which included data up to December 2021.^17^ Using contemporaneous data in OpenSAFELY, our study demonstrates that, while incidence rates for COPD and asthma have gradually recovered following these initial decreases, large deficits remained as of November 2024, suggesting many cases remain undiagnosed as a consequence of the pandemic. An important contributing factor is likely to be the sudden disruption to spirometry and lung function testing during the pandemic, resulting in large backlogs in testing.^21^ As such, equitable access to spirometry has been identified as a priority area for the NHS as it recovers from the pandemic.^21^

Of all conditions studied, the largest relative and absolute deficits in diagnoses were observed for depression. After an initial decrease in diagnoses during the early pandemic, the incidence of depression partly recovered by late 2021, but has declined markedly since 2022. While a disproportionate impact of the early pandemic on mental health diagnoses has been reported previously,^17,19^ the recent fall in new diagnoses of depression warrants further investigation. Our findings are supported by decreases in new initiations of antidepressant medications and referrals to mental health services for depression in England since 2022, following large increases previously.^22,23^ It is important to note, however, that the observed decline in new diagnoses may not necessarily equate to a decrease in the underlying incidence of depression. New claimants of disability-related benefits due to mental health conditions have increased by 190% in England between 2019/20 and 2023/24.^24^ One possible explanation for this disconnect is that people with symptoms of depression are taking longer to be formally diagnosed due to increasing pressures on the NHS.^25,26^ Primary care services have faced particular challenges with increased demand since the pandemic. Although the total number of face-to-face consultations has increased, the proportion of remote consultations has also grown, which together with pressures on time in consultations, may make it more challenging to identify non-verbal cues associated with depression.^27^ It is also possible that individuals with symptoms of depression may be accessing services without having their diagnoses recorded in primary or secondary care records. Following a national drive to expand access to psychological therapies, referrals to NHS Talking Therapies services increased by nearly two-thirds between 2013/14 and 2023/24 (from 1.1 million to 1.8 million referrals, respectively), with self-referrals (i.e. without necessarily needing to consult a healthcare professional prior to referral) constituting 69% of all referrals.^28^ Recommendations from the National Institute for Health and Care Excellence (NICE) – updated in June 2022 – emphasised the importance of therapy-based treatments over antidepressants for new cases of less severe depression.^29^ This, in turn, may have contributed to a change in how clinicians are recording depression diagnoses. In our sensitivity analysis that included additional diagnostic codes for symptoms of depression (e.g. depressed mood), the relative deficit in depression diagnoses was attenuated; indicating a shift towards recording symptom-related codes over formal depression diagnoses. Previous studies reported similar findings after the introduction of the Quality and Outcomes Framework (QOF) incentivisation scheme in the NHS, which resulted in GPs switching from recording diagnostic codes for depression to symptom-related codes.^30^

A large, persistent deficit in diagnoses was also observed for osteoporosis. Delays in diagnosis and the initiation of bone protection therapy can substantially impact prognosis for individuals with osteoporosis. Osteoporotic fractures are associated with a high burden of disability and functional impairment,^31^ with estimates of mortality in the first year after hip fracture ranging from 15% to 36%.^32^ Bisphosphonates reduce the risk of hip and vertebral fractures in post-menopausal women by 35% and 54%, respectively;^33^ highlighting the importance of early diagnosis and treatment. Our findings therefore suggest that a national case-finding strategy may be warranted to identify individuals who have not yet been diagnosed with or treated for osteoporosis as a consequence of the pandemic.

In contrast, a large increase in newly-recorded diagnoses of CKD has been evident since 2022. While this may partially represent a catch-up in diagnoses following service disruption during the pandemic, the observed increase in CKD diagnoses was 33% higher than expected based upon pre-pandemic trends. Subgroup analyses highlighted that much of this increase occurred in individuals aged 80 years and over, and in those from less deprived socioeconomic quintiles. There are numerous factors that could have potentially contributed to this increase, including accelerated GFR decline following COVID-19 infection,^34,35^ the inclusion of CKD as a risk factor in national cardiovascular prevention audits (e.g. CVDPREVENT),^36^ initiatives to facilitate the use of novel therapies to prevent CKD progression (e.g. SGLT-2 inhibitors),^37^ and remote albuminuria testing programmes.^38^ In August 2021, updated guidance on CKD assessment and management was published by NICE, containing recommendations for the investigation of CKD, including screening of at-risk individuals, which may have led to enhanced case detection after its implementation.^39^ Additionally, updated NICE guidelines recommended removal of ethnicity-related adjustment factors when estimating glomerular filtration rates (eGFR). Use of ethnicity-based eGFR was demonstrated to overestimate GFR and under-recognise CKD in individuals of Black ethnicity in the UK,^39,40^ which may have contributed to the observed increase in individuals of Black ethnicity meeting criteria for CKD diagnosis following its removal. However, the comparable relative increase in CKD diagnoses among individuals of White ethnicity suggests that this factor alone is unlikely to explain the large overall increase in CKD diagnoses.

Importantly, the methodology utilised in this study provides a mechanism through which to highlight variation by age, sex, ethnicity, and socioeconomic status. Despite England having a universal, free at the point-of-access healthcare system, we observed compensatory post-pandemic increases in dementia diagnoses for individuals of White ethnicity and those from less deprived socioeconomic quintiles, whereas diagnosis rates remained lower than expected among individuals from other ethnic backgrounds and more deprived communities. By highlighting and exploring these disparities in more detail – for example, through more granular analyses by ethnicity and region – these data could inform the development and targeting of quality improvement interventions towards reducing inequity. Contemporaneous data sources in OpenSAFELY provide opportunities not only for surveillance and early detection of diagnostic trends and inequity, but also the ability to monitor the impact of quality improvement interventions, while minimising the burden of manual data collection and maximising the inclusion of populations who are often under-represented in clinical studies.^41-43^ Although the primary focus of our study was to describe changes in the incidence and prevalence of long-term conditions, the methodology and analysis code can be readily adapted to assess care quality benchmarked against clinical guidelines,^6,7^ and evaluate the impact of strategies aimed at improving healthcare delivery.^41^

Our study had several strengths, including the use of nationally representative data from over 27 million individuals to provide updated estimates of incidence and prevalence for 19 long-term conditions, spanning multiple disease areas. This approach can be expanded to assess the incidence, prevalence, and management of numerous other diseases at national, regional and local levels, including rare diseases with limited epidemiological data. Additionally, all analyses can updated regularly and made publicly available,^44^ enabling near real-time monitoring of changing incidence rates – for example, in response to the implementation of clinical guidelines.

There are also limitations that must be acknowledged. As with other studies utilising coded health data, there is a potential for diagnostic misclassification. While these data can be used to highlight temporal trends in new diagnoses recorded in primary and secondary care, we were unable to infer whether the observed differences represented true changes in underlying disease incidence (e.g. following SARS-CoV-2 infection), changes in the recording of diagnoses (e.g. due to updated guidelines and/or incentivisation), changes in diagnostic testing (e.g. increased testing of B-type natriuretic peptide for heart failure), or changes in service delivery (e.g. care provision outside of primary and secondary care settings). Trends in diagnoses must be considered against the backdrop of abrupt changes in health care utilisation during the pandemic: the number of general practice appointments in England fell by one third between March and April 2020, before increasing above pre-pandemic levels since then.^45^ To maximise case capture, we utilised data from both primary care and secondary care sources; however, it is important to acknowledge the differences between diagnoses made in primary care and hospital settings. We were unable to age and sex-standardise our subgroup analyses by socioeconomic status or ethnicity due to the computationally-intensive nature of these analyses; although crude and adjusted estimates for the overall cohort were comparable, suggesting this is unlikely to have meaningfully altered our findings. We reported incidence trends separated into five ethnicity categories for most, but not all, conditions, due to small numbers of incident cases and the potential for disclosure; this also precluded more granular reporting of differences within these broad ethnicity categories. Importantly, as these analyses were conducted using routinely collected data from the NHS of England, they may not necessarily be generalisable to other healthcare systems or settings.

In conclusion, our study utilised routinely collected health data for 27 million individuals to provide a comprehensive epidemiological analysis of chronic diseases in England. We highlighted large post-pandemic deficits in diagnoses for conditions including depression, COPD, asthma and osteoporosis, contrasting a surge in recorded CKD diagnoses since the pandemic. Importantly, this study demonstrates the potential for real-time health data to transform disease monitoring, identify inequities across under-served populations, and inform strategies to improve case detection and reduce diagnostic delays.

## Acknowledgments

We are very grateful for all the support received from the TPP Technical Operations team throughout this work, and for generous assistance from the information governance and database teams at NHS England and the NHS England Transformation Directorate.

## Data availability and sharing

All data were linked, stored and analysed securely using the OpenSAFELY platform, https://www.opensafely.org/, as part of the NHS England OpenSAFELY COVID-19 service. Data include pseudonymised data such as coded diagnoses, medications and physiological parameters. No free text data are included. No GP data from patients who have registered a Type-1 opt-out with their GP surgery were included in this study. All code is shared openly for review and re-use under MIT open license https://github.com/opensafely/disease_incidence. Detailed pseudonymised patient data is potentially re-identifiable and therefore not shared.

Access to the underlying identifiable and potentially re-identifiable pseudonymised electronic health record data is tightly governed by various legislative and regulatory frameworks, and restricted by best practice. The data in the NHS England OpenSAFELY COVID-19 service is drawn from General Practice data across England where TPP is the data processor. TPP developers initiate an automated process to create pseudonymised records in the core OpenSAFELY database, which are copies of key structured data tables in the identifiable records. These pseudonymised records are linked onto key external data resources that have also been pseudonymised via SHA-512 one-way hashing of NHS numbers using a shared salt. University of Oxford, Bennett Institute for Applied Data Science developers and PIs, who hold contracts with NHS England, have access to the OpenSAFELY pseudonymised data tables to develop the OpenSAFELY tools. These tools in turn enable researchers with OpenSAFELY data access agreements to write and execute code for data management and data analysis without direct access to the underlying raw pseudonymised patient data, and to review the outputs of this code. All code for the full data management pipeline — from raw data to completed results for this analysis — and for the OpenSAFELY platform as a whole is available for review at github.com/OpenSAFELY. The data management and analysis code for this paper was led by MDR and contributed to by JBG.

## Information governance

NHS England is the data controller of the NHS England OpenSAFELY COVID-19 Service; TPP is the data processor; all study authors using OpenSAFELY have the approval of NHS England.^46^ This implementation of OpenSAFELY is hosted within the TPP environment, which is accredited to the ISO 27001 information security standard and is NHS IG Toolkit compliant^47^. Patient data has been pseudonymised for analysis and linkage using industry standard cryptographic hashing techniques; all pseudonymised datasets transmitted for linkage onto OpenSAFELY are encrypted; access to the NHS England OpenSAFELY COVID-19 service is via a virtual private network (VPN) connection; the researchers hold contracts with NHS England and only access the platform to initiate database queries and statistical models; all database activity is logged; only aggregate statistical outputs leave the platform environment following best practice for anonymisation of results such as statistical disclosure control for low cell counts^48^. The service adheres to the obligations of the UK General Data Protection Regulation (UK GDPR) and the Data Protection Act 2018. The service previously operated under notices initially issued in February 2020 by the Secretary of State under Regulation 3(4) of the Health Service (Control of Patient Information) Regulations 2002 (COPI Regulations), which required organisations to process confidential patient information for COVID-19 purposes; this set aside the requirement for patient consent^48^. As of 1 July 2023, the Secretary of State has requested that NHS England continue to operate the Service under the COVID-19 Directions 2020^50^. In some cases of data sharing, the common law duty of confidence is met using, for example, patient consent or support from the Health Research Authority Confidentiality Advisory Group^51^. Taken together, these provide the legal bases to link patient datasets using the service. GP practices, which provide access to the primary care data, are required to share relevant health information to support the public health response to the pandemic, and have been informed of how the service operates. This study was supported by Dr Elizabeth Price (National Clinical Lead for National Early Inflammatory Arthritis Audit) as senior sponsor, and approval to undertake this study under the remit of service evaluation was obtained from King’s College Hospital NHS Foundation Trust.

## Contributorship statement

Contributions are as follows: Conceptualisation: MDR, JBG; Methodology: MDR, JBG, SN, KB, MG, ZJ, AS, BMK, RH, JM: Data curation: JBG, MDR, AS, JM, RH, BMK, PI, SB, AM, BG; Formal analysis: MDR, JBG, SN; Interpretation of findings: all authors; Writing – original draft: MDR; Writing – revising, review and editing: all authors. All authors read and approved the final manuscript. MDR and JBG are the guarantors for the article, and accept full responsibility for the work and/or the conduct of the study, had access to the data, and controlled the decision to publish. MDR and JBG directly accessed and verified the underlying data reported in the manuscript. The corresponding author attests that all listed authors meet authorship criteria and that no others meeting the criteria have been omitted.

The Corresponding Author has the right to grant on behalf of all authors and does grant on behalf of all authors, a worldwide licence to the Publishers and its licensees in perpetuity, in all forms, formats and media (whether known now or created in the future), to i) publish, reproduce, distribute, display and store the Contribution, ii) translate the Contribution into other languages, create adaptations, reprints, include within collections and create summaries, extracts and/or, abstracts of the Contribution, iii) create any other derivative work(s) based on the Contribution, iv) to exploit all subsidiary rights in the Contribution, v) the inclusion of electronic links from the Contribution to third party material where-ever it may be located; and, vi) licence any third party to do any or all of the above.

## Transparency statement

The lead author (the manuscript’s guarantor) affirms that the manuscript is an honest, accurate, and transparent account of the study being reported; that no important aspects of the study have been omitted; and that any discrepancies from the study as originally planned have been explained.

