## Supplementary Appendix for "Time trends in new diagnoses of 19 long-term conditions: a population-level cohort study in England using OpenSAFELY"

### **Supplementary Figure S1**. Monthly incidence rates for 19 long-term conditions in England between April 1, 2016, to November 30, 2024.

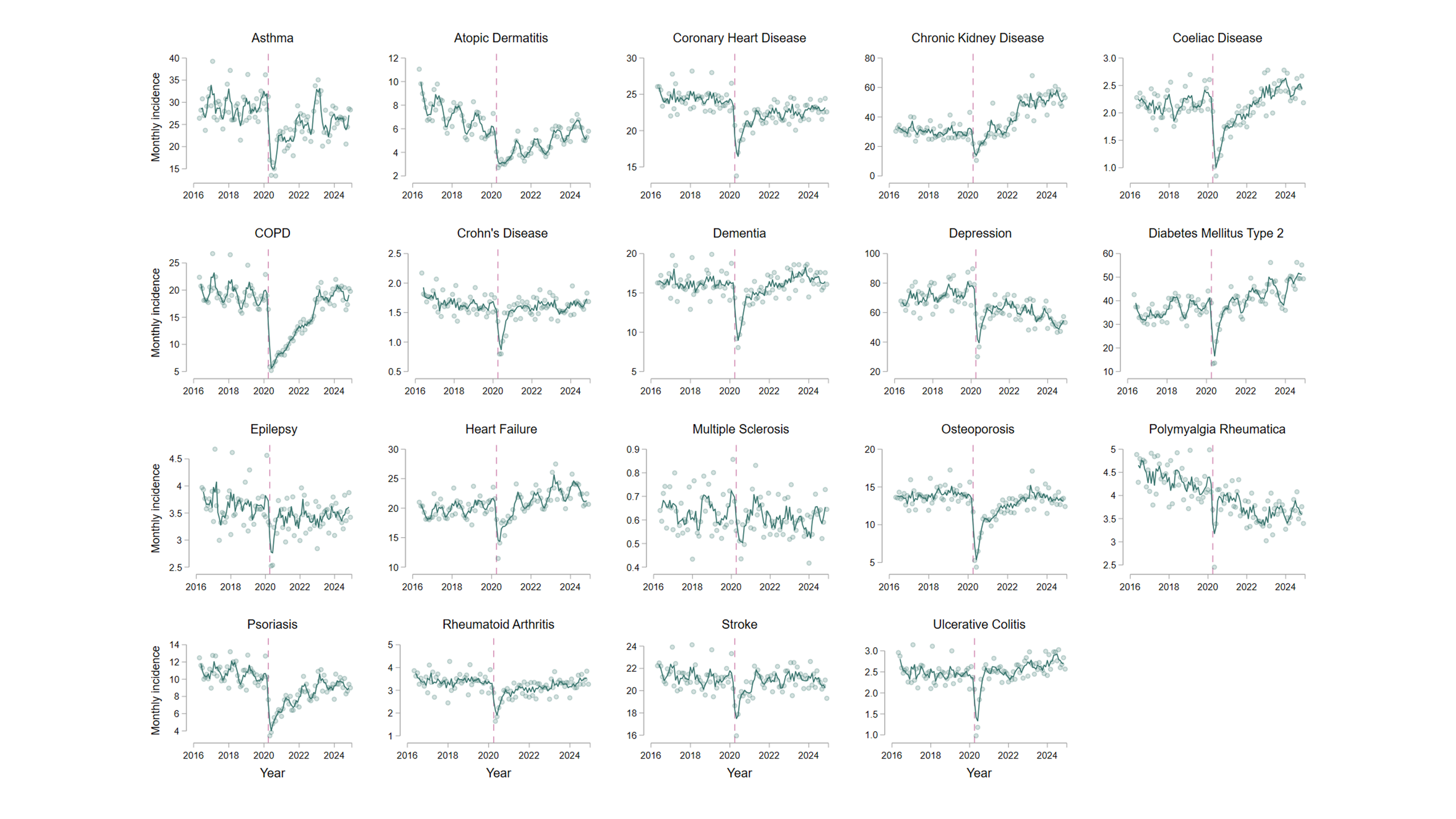
Age and sex-adjusted incidence rates are shown per 100,000 population. Individual data points represent the monthly incidence for each condition, and are presented alongside 3-monthly rolling averages of the current, preceding and subsequent months. The vertical dashed line corresponds to the onset of the first COVID-19 lockdown in England (March 2020).

### **Supplementary Figure S2**. Comparison between crude and adjusted incidence rates for 19 long-term conditions in England between April 1, 2016, to November 30, 2024.

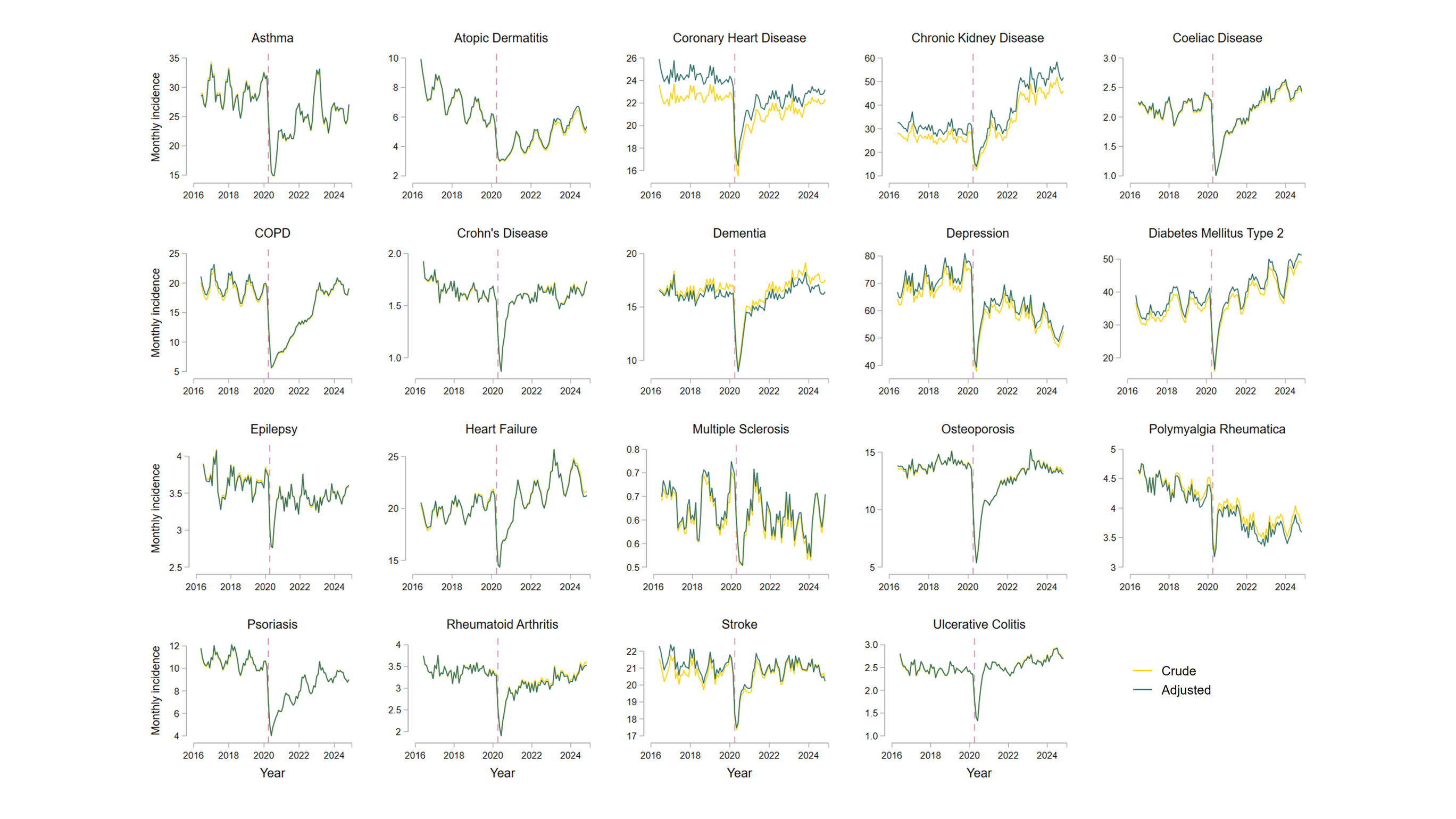
Unadjusted (crude) and age and sex-adjusted incidence rates are shown per 100,000 population as 3-monthly rolling averages of the current, preceding and subsequent months. The vertical dashed line corresponds to the onset of the first COVID-19 lockdown in England (March 2020).

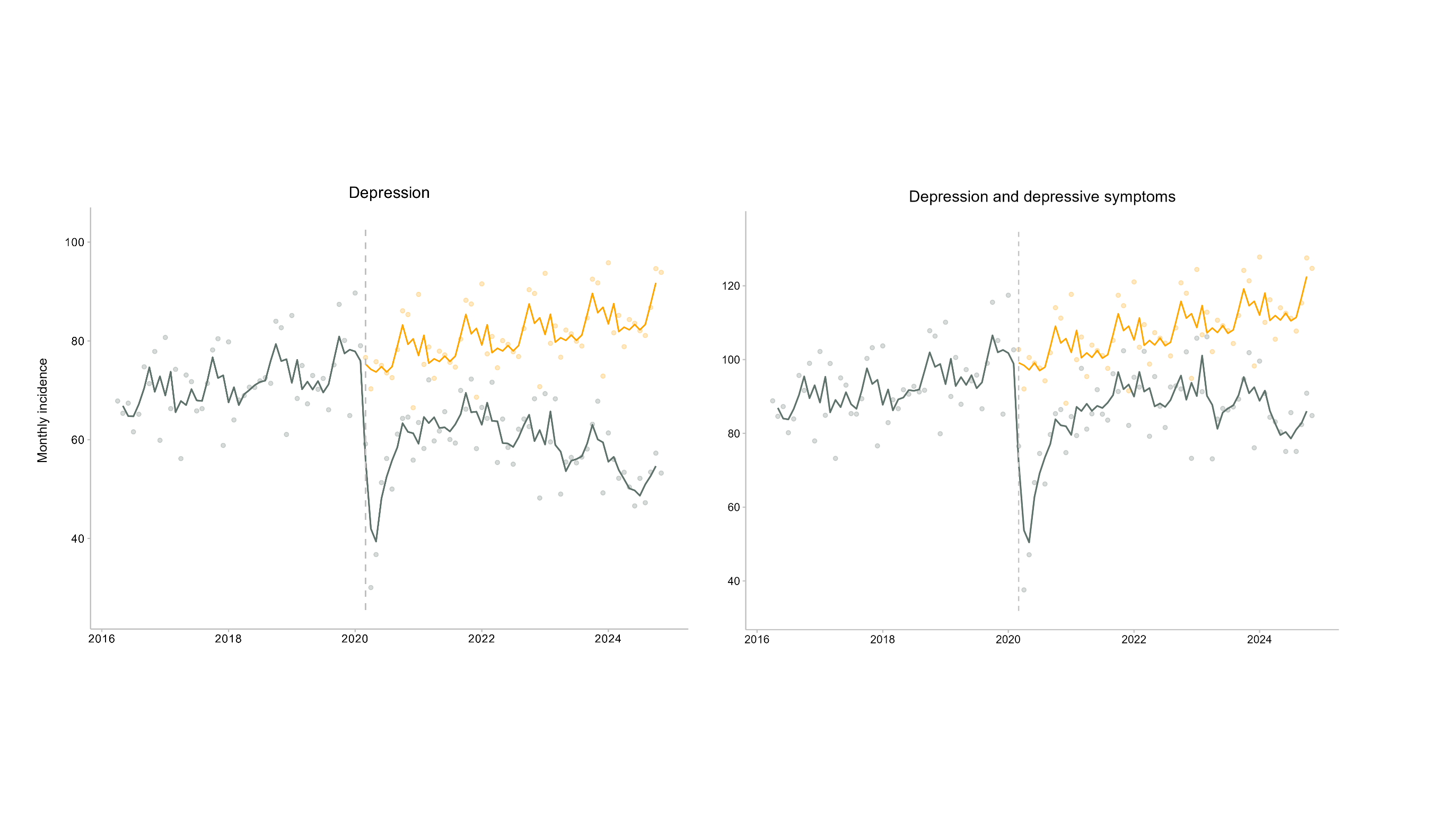

### **Supplementary Figure S3**. Monthly incidence rates for depression diagnoses, comparing trends with and without the inclusion of additional diagnostic codes for depressive symptoms.

In the left panel, the observed (dark green) and expected (orange) incidence rates for depression are compared after the onset of the COVID-19 pandemic in England. In the right panel, the observed and expected incidence rates for depression are compared when using an expanded depression codelist that included symptomatic codes suggestive of depression (e.g. depressed mood). Expected incidence rates after March 2020 (vertical dashed line) were estimated using seasonal autoregressive integrated moving averages (SARIMA) models, utilising data from April 1, 2016, to February 28, 2020. Individual data points, representing age and sex-adjusted monthly incidence rates per 100,000 population, are presented alongside 3-monthly rolling averages of the current, preceding and subsequent months.

### **Supplementary Figure S4**. Monthly incidence rates by age band for 19 long-term conditions in England between April 1, 2016, to November 30, 2024.

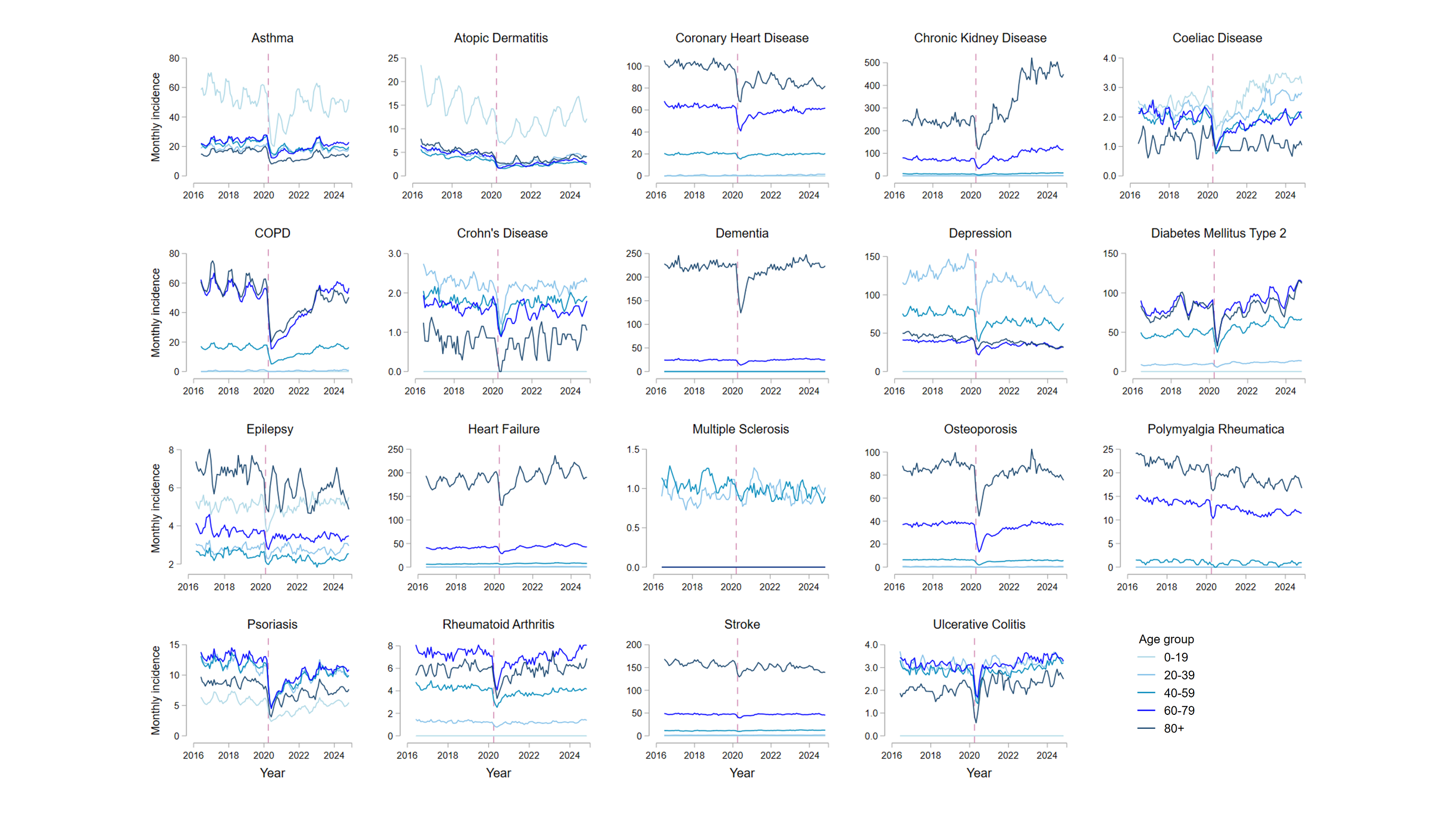
Monthly incidence rates for each condition are shown by age band (0-19, 20-39, 40-59, 60-79 and 80+ years), and presented as 3-monthly rolling averages of the current, preceding and subsequent months per 100,000 population. The vertical dashed line corresponds to the onset of the first COVID-19 lockdown in England (March 2020).

### **Supplementary Figure S5**. Monthly incidence rates by ethnicity for 11 long-term conditions in England between April 1, 2016, to November 30, 2024.

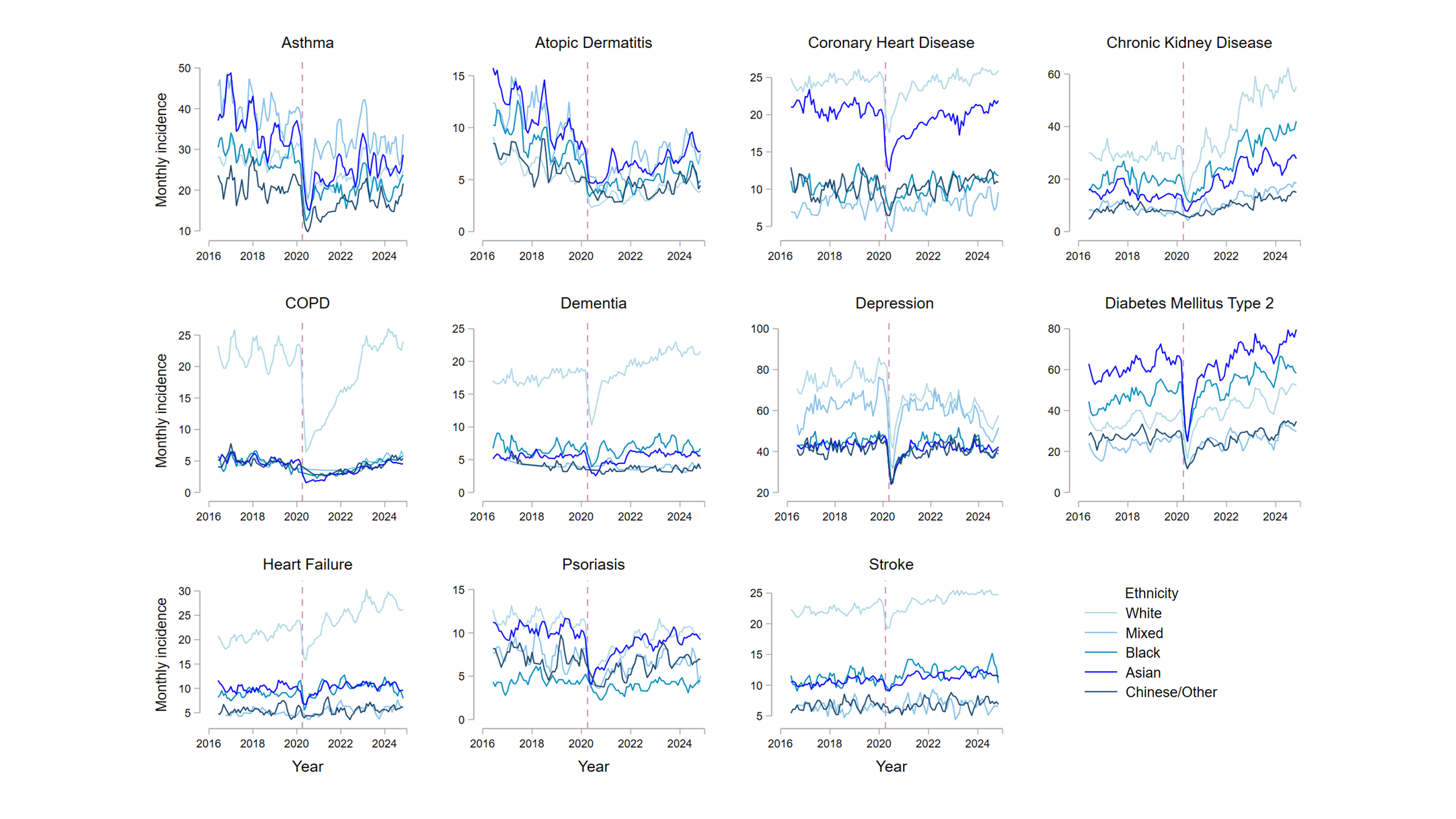

Monthly incidence rates for each condition are shown by ethnicity (categorised into White, Mixed, Asian or Asian British, Black or Black British, or Chinese/Other ethnic groups), and presented as 3-monthly rolling averages of the current, preceding and subsequent months per 100,000 population. The vertical dashed line corresponds to the onset of the first COVID-19 lockdown in England (March 2020). No age or sex-standardisation was performed due to computational requirements. Conditions with small numbers of incident diagnoses when analysed separately by ethnicity are not shown, due to the potential for disclosure.

### **Supplementary Figure S6**. Annual prevalence for 19 long-term conditions in England between April 1, 2016, and March 31, 2023.

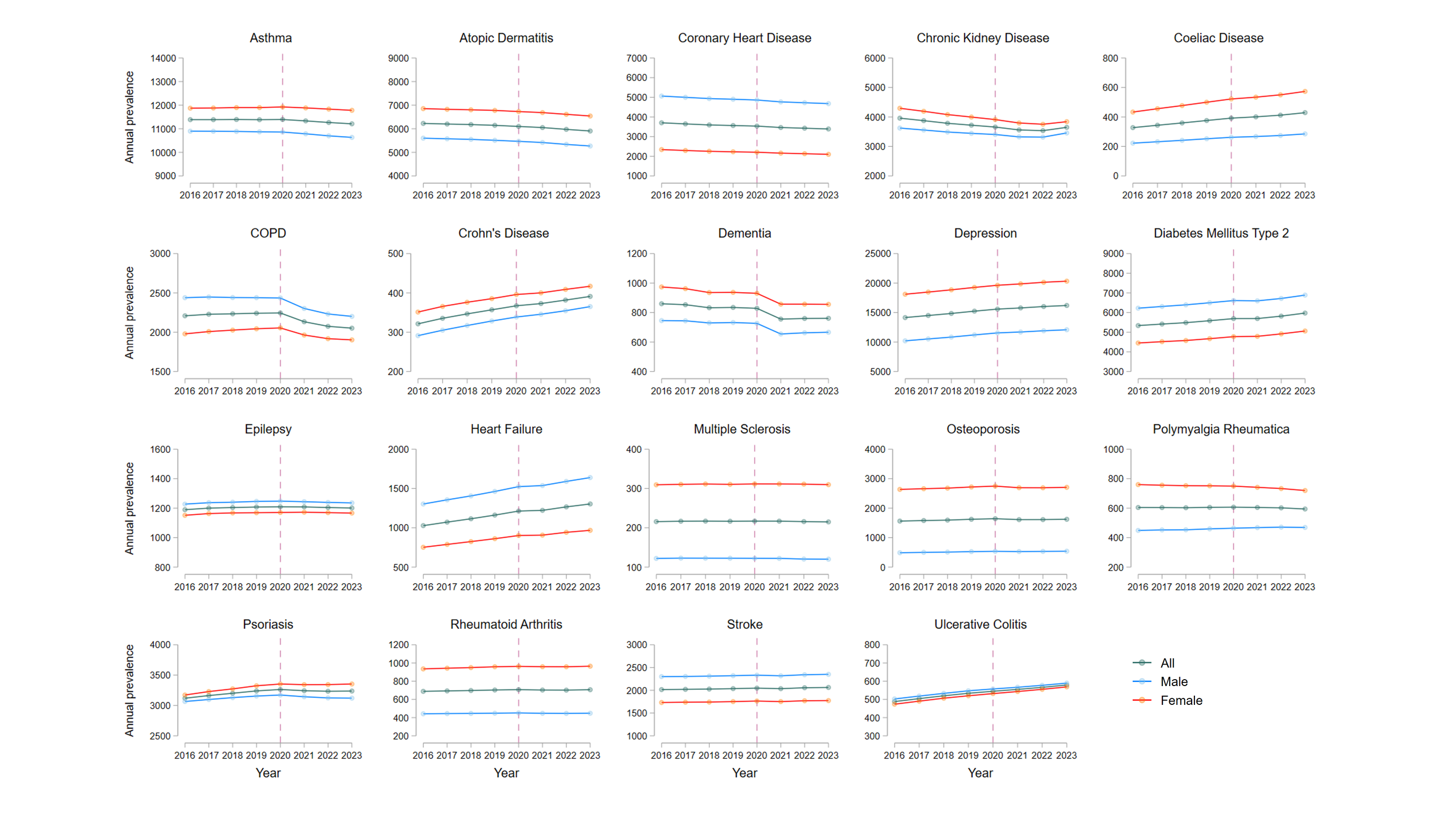

Annual prevalence per 100,000 population is shown overall (age and sex-adjusted) and separately by sex (age-adjusted). The vertical dashed line corresponds to the onset of the first COVID-19 lockdown in England (March 2020).

### **Supplementary Figure S7**. Comparison between crude and age and sex-adjusted prevalence for 19 long-term conditions in England between April 1, 2016, and March 31, 2023.

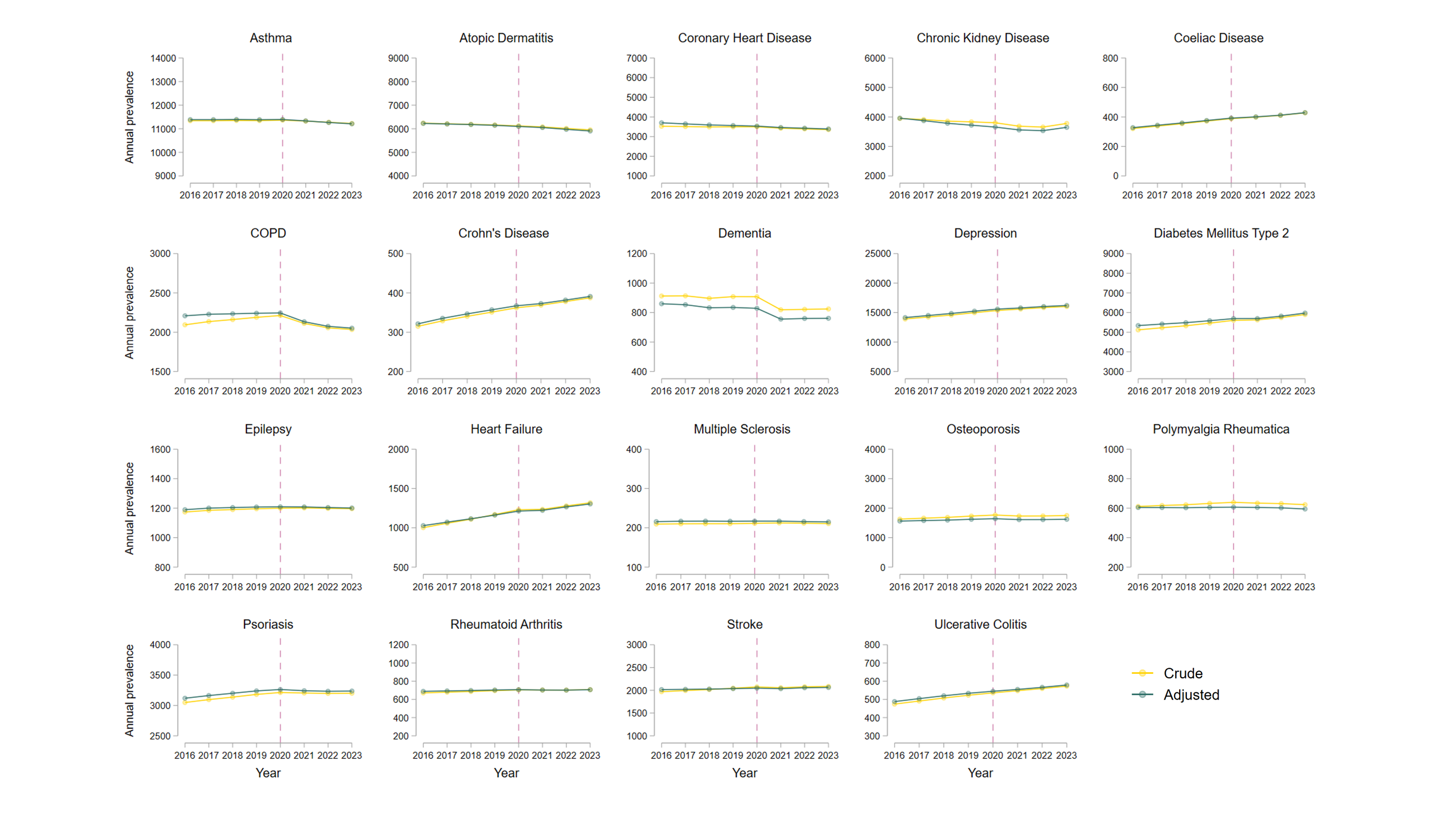
Unadjusted (crude) and age and sex-adjusted annual prevalence per 100,000 population are shown. The vertical dashed line corresponds to the onset of the first COVID-19 lockdown in England (March 2020).

### **Supplementary Table S1.** Sociodemographic characteristics for the reference population and for individuals with incident diagnoses of 19 long-term health conditions.

|  | **Reference**  **population** | **Asthma** | **Atopic dermatitis** | **Coronary heart disease** | **Chronic kidney disease** | **Coeliac disease** | **COPD** | **Crohn’s disease** | **Dementia** | **Depression** |
| --- | --- | --- | --- | --- | --- | --- | --- | --- | --- | --- |
|  | **n=27,132,190** | **n=453,700** | **n=92,150** | **n=436,660** | **n=637,955** | **n=42,900** | **n=334,545** | **n=33,575** | **n=336,030** | **n=1,120,320** |
| Mean (SD) age, years | 42.1 (23.1) | 35.3 (24.7) | 34 (25.9) | 67.3 (13.0) | 74.3 (12.2) | 38.8 (22.1) | 66.6 (12.4) | 42.5 (19.5) | 82.1 (8.2) | 38.9 (18.1) |
| Sex |  |  |  |  |  |  |  |  |  |  |
| Female | 13,625,625 (50.2%) | 249,825 (55.1%) | 51,825 (56.2%) | 164,900 (37.8%) | 332,220 (52.1%) | 28,005 (65.3%) | 159,625 (47.7%) | 16,930 (50.4%) | 199,470 (59.4%) | 637,510 (56.9%) |
| Male | 13,506,560 (49.8%) | 203,875 (44.9%) | 40,330 (43.8%) | 271,760 (62.2%) | 305,735 (47.9%) | 14,900 (34.7%) | 174,920 (52.3%) | 16,645 (49.6%) | 136,560 (40.6%) | 482,810 (43.1%) |
| Ethnicity |  |  |  |  |  |  |  |  |  |  |
| White | 18,093,920 (66.7%) | 307,985 (67.9%) | 59,300 (64.4%) | 335,920 (76.9%) | 501,835 (78.7%) | 31,715 (73.9%) | 277,375 (82.9%) | 24,980 (74.4%) | 260,570 (77.5%) | 821,710 (73.3%) |
| Asian/Asian British | 2,225,040 (8.2%) | 35,090 (7.7%) | 9,645 (10.5%) | 26,405 (6.0%) | 25,250 (4.0%) | 3,135 (7.3%) | 5,560 (1.7%) | 1,990 (5.9%) | 7,310 (2.2%) | 53,425 (4.8%) |
| Black/Black British | 849,460 (3.1%) | 9,325 (2.1%) | 2,370 (2.6%) | 4,680 (1.1%) | 11,440 (1.8%) | 225 (0.5%) | 1,910 (0.6%) | 360 (1.1%) | 3,010 (0.9%) | 18,260 (1.6%) |
| Mixed | 441,905 (1.6%) | 8,010 (1.8%) | 1,735 (1.9%) | 2,230 (0.5%) | 3,125 (0.5%) | 310 (0.7%) | 1,105 (0.3%) | 400 (1.2%) | 940 (0.3%) | 15,425 (1.4%) |
| Chinese or other ethnicity | 748,670 (2.8%) | 5,925 (1.3%) | 1,525 (1.7%) | 3,680 (0.8%) | 3,395 (0.5%) | 305 (0.7%) | 1,425 (0.4%) | 350 (1.0%) | 1,225 (0.4%) | 13,730 (1.2%) |
| Unknown | 4,773,200 (17.6%) | 87,365 (19.3%) | 17,570 (19.1%) | 63,745 (14.6%) | 92,910 (14.6%) | 7,205 (16.8%) | 47,175 (14.1%) | 5,490 (16.4%) | 62,975 (18.7%) | 197,775 (17.7%) |
| Index of multiple deprivation |  |  |  |  |  |  |  |  |  |  |
| 1 most deprived | 5,287,175 (19.5%) | 99,930 (22.0%) | 19,980 (21.7%) | 80,635 (18.5%) | 108,255 (17%) | 7,110 (16.6%) | 92,500 (27.6%) | 6,420 (19.1%) | 56,935 (16.9%) | 266,620 (23.8%) |
| 2 | 5,311,040 (19.6%) | 88,275 (19.5%) | 17,610 (19.1%) | 83,840 (19.2%) | 118,665 (18.6%) | 7,300 (17.0%) | 72,455 (21.7%) | 6,440 (19.2%) | 62,730 (18.7%) | 227,970 (20.3%) |
| 3 | 5,633,610 (20.8%) | 90,695 (20.0%) | 18,300 (19.9%) | 94,830 (21.7%) | 140,780 (22.1%) | 8,790 (20.5%) | 67,720 (20.2%) | 6,915 (20.6%) | 75,005 (22.3%) | 220,745 (19.7%) |
| 4 | 5,174,070 (19.1%) | 81,795 (18.0%) | 16,855 (18.3%) | 87,660 (20.1%) | 133,915 (21.0%) | 8,950 (20.9%) | 54,440 (16.3%) | 6,535 (19.5%) | 70,985 (21.1%) | 193,760 (17.3%) |
| 5 least deprived | 4,690,345 (17.3%) | 75,505 (16.6%) | 16,065 (17.4%) | 80,345 (18.4%) | 123,240 (19.3%) | 9,075 (21.2%) | 40,810 (12.2%) | 5,980 (17.8%) | 64,870 (19.3%) | 165,315 (14.8%) |
| Unknown | 1,035,945 (3.8%) | 17,495 (3.9%) | 3,345 (3.6%) | 9,350 (2.1%) | 13,100 (2.1%) | 1,675 (3.9%) | 6,625 (2.0%) | 1,285 (3.8%) | 5,500 (1.6%) | 45,910 (4.1%) |

**Supplementary Table S1 (continued).** Sociodemographic characteristics for the reference population and for or individuals with incident diagnoses of 19 long-term health conditions.

|  | **Diabetes mellitus type 2** | **Epilepsy** | **Heart failure** | **Multiple sclerosis** | **Osteoporosis** | **Polymyalgia rheumatica** | **Psoriasis** | **Rheumatoid arthritis** | **Stroke/TIA** | **Ulcerative colitis** |
| --- | --- | --- | --- | --- | --- | --- | --- | --- | --- | --- |
|  | **n=737,745** | **n=69,500** | **n=423,515** | **n=12,780** | **n=265,805** | **n=83,995** | **n=187,850** | **n=67,695** | **n=420,360** | **n=52,340** |
| Mean (SD) age, years | 60.2 (14.6) | 42.4 (26.5) | 75.8 (13.2) | 43.3 (13.8) | 72.4 (12.3) | 72.6 (9.9) | 45.5 (20.6) | 59.4 (16.4) | 72.3 (14.3) | 47.3 (19.4) |
| Sex |  |  |  |  |  |  |  |  |  |  |
| Female | 330,125 (44.7%) | 32,325 (46.5%) | 190,585 (45.0%) | 8,790 (68.8%) | 213,360 (80.3%) | 50,005 (59.5%) | 100,125 (53.3%) | 44,715 (66.1%) | 205,555 (48.9%) | 25,830 (49.4%) |
| Male | 407,620 (55.3%) | 37,175 (53.5%) | 232,930 (55.0%) | 3,990 (31.2%) | 52,445 (19.7%) | 33,990 (40.5%) | 87,725 (46.7%) | 22,980 (33.9%) | 214,805 (51.1%) | 26,510 (50.6%) |
| Ethnicity |  |  |  |  |  |  |  |  |  |  |
| White | 526,730 (71.4%) | 49,415 (71.1%) | 325,790 (76.9%) | 9,900 (77.4%) | 216,130 (81.3%) | 70,955 (84.5%) | 140,225 (74.6%) | 52,310 (77.3%) | 323,735 (77.0%) | 39,635 (75.7%) |
| Asian/Asian British | 78,445 (10.6%) | 3,455 (5.0%) | 13,670 (3.2%) | 545 (4.3%) | 7,920 (3.0%) | 1,300 (1.5%) | 12,370 (6.6%) | 4,715 (7.0%) | 14,860 (3.5%) | 3,100 (5.9%) |
| Black/Black British | 21,500 (2.9%) | 1,435 (2.1%) | 4,285 (1.0%) | 280 (2.2%) | 1,435 (0.5%) | 470 (0.6%) | 1,835 (1.0%) | 1,120 (1.7%) | 5,195 (1.2%) | 560 (1.1%) |
| Mixed | 6,840 (0.9%) | 905 (1.3%) | 1,470 (0.3%) | 170 (1.3%) | 825 (0.3%) | 200 (0.2%) | 1,865 (1.0%) | 525 (0.8%) | 1,860 (0.4%) | 520 (1%) |
| Chinese or other ethnicity | 9,445 (1.3%) | 645 (0.9%) | 1,960 (0.5%) | 145 (1.1%) | 1,855 (0.7%) | 260 (0.3%) | 2,305 (1.2%) | 630 (0.9%) | 2,405 (0.6%) | 490 (0.9%) |
| Unknown | 94,780 (12.8%) | 13,645 (19.6%) | 76,335 (18.0%) | 1,745 (13.6%) | 37,635 (14.2%) | 10,805 (12.9%) | 29,245 (15.6%) | 8,400 (12.4%) | 72,300 (17.2%) | 8,030 (15.3%) |
| Index of multiple deprivation |  |  |  |  |  |  |  |  |  |  |
| 1 most deprived | 171,475 (23.2%) | 17,215 (24.8%) | 78,030 (18.4%) | 2,140 (16.7%) | 41,045 (15.4%) | 8,790 (10.5%) | 38,685 (20.6%) | 12,655 (18.7%) | 74,540 (17.7%) | 8,890 (17%) |
| 2 | 155,615 (21.1%) | 14,000 (20.1%) | 81,260 (19.2%) | 2,440 (19.1%) | 46,720 (17.6%) | 13,095 (15.6%) | 36,090 (19.2%) | 13,230 (19.5%) | 78,840 (18.8%) | 9,755 (18.6%) |
| 3 | 153,440 (20.8%) | 13,700 (19.7%) | 92,795 (21.9%) | 2,660 (20.8%) | 58,030 (21.8%) | 19,720 (23.5%) | 38,325 (20.4%) | 14,790 (21.8%) | 91,685 (21.8%) | 11,025 (21.1%) |
| 4 | 131,035 (17.8%) | 11,740 (16.9%) | 86,580 (20.4%) | 2,590 (20.3%) | 57,680 (21.7%) | 20,230 (24.1%) | 35,735 (19.0%) | 13,305 (19.7%) | 86,815 (20.7%) | 10,565 (20.2%) |
| 5 least deprived | 107,810 (14.6%) | 10,650 (15.3%) | 77,310 (18.3%) | 2,435 (19.0%) | 56,395 (21.2%) | 20,570 (24.5%) | 32,560 (17.3%) | 11,905 (17.6%) | 80,255 (19.1%) | 10,100 (19.3%) |
| Unknown | 18,375 (2.5%) | 2,195 (3.2%) | 7,540 (1.8%) | 520 (4.1%) | 5,935 (2.2%) | 1,590 (1.9%) | 6,460 (3.4%) | 1,810 (2.7%) | 8,225 (2.0%) | 2,000 (3.8%) |

Sociodemographic characteristics are summarised for the reference population at the study midpoint (August 2020), and at the time of diagnosis for individuals from the reference population who had incident diagnoses of one or more of 19 long-term conditions during the study period (April 1, 2016, to November 30, 2024). Counts have been rounded to the nearest 5, to reduce the risk of disclosure. As such, column totals may differ from the sum of the individual variables. COPD: chronic obstructive pulmonary disease; TIA: transient ischaemic attack.

### **Supplementary Table S2.** Differences between expected and observed incidence rates for 19 long-term conditions after the onset of the COVID-19 pandemic in England, separated by year.

|  | **March 2020 to February 2021** | | **March 2021 to February 2022** | | **March 2022 to February 2023** | | **March 2023 to November 2024** | |
| --- | --- | --- | --- | --- | --- | --- | --- | --- |
| **Condition** | Absolute difference between expected and observed IR per 100,000 population (95% CI) | Relative difference between expected and observed IR per 100,000 population (95% CI) | Absolute difference between expected and observed IR per 100,000 population (95% CI) | Relative difference between expected and observed IR per 100,000 population (95% CI) | Absolute difference between expected and observed IR per 100,000 population (95% CI) | Relative difference between expected and observed IR per 100,000 population (95% CI) | Absolute difference between expected and observed IR per 100,000 population (95% CI) | Relative difference between expected and observed IR per 100,000 population (95% CI) |
| Asthma | -110.8  (-122.8, -98.9) | -31.8%  (-34.0, -29.3) | -66.6  (-79.6, -53.5) | -19.1%  (-22.0, -15.9) | -20.6  (-34.7, -6.47) | -5.90%  (-9.55, -1.93) | -66.2  (-86.7, -45.8) | -10.9%  (-13.9, -7.83) |
| Atopic Dermatitis | -26.4  (-31.5, -21.2) | -39.1%  (-43.4, -34.0) | -9.73  (-18.4, -1.08) | -16.0%  (-26.4, -2.06) | 1.40  (-11.2, 14.0) | 2.58%  (-16.7, 33.6) | 42.5  (17.1, 67.9) | 54.1%  (16.5, 127.7) |
| Coronary Heart Disease | -52.7  (-58.6, -46.8) | -18.4%  (-20.0, -16.6) | -18.0  (-24.5, -11.5) | -6.33%  (-8.43, -4.13) | -11.7  (-19.6, -3.77) | -4.14%  (-6.76, -1.38) | -7.50  (-19.5, 4.45) | -1.54%  (-3.91, 0.94) |
| Chronic Kidney Disease | -92.3  (-116.2, -68.4) | -25.4%  (-30.0, -20.1) | 23.3  (-15.7, 62.2) | 6.33%  (-3.87, 19.0) | 181.9  (126.7, 237.0) | 49.1%  (29.8, 75.2) | 458.7  (351.0, 566.4) | 70.7%  (46.4, 104.7) |
| Coeliac Disease | -8.34  (-9.64, -7.05) | -32.0%  (-35.2, -28.4) | -3.00  (-4.32, -1.66) | -11.5%  (-15.7, -6.69) | 1.52  (0.16, 2.89) | 5.83%  (0.57, 11.67) | 5.71  (3.84, 7.58) | 12.5%  (8.1, 17.4) |
| COPD | -121.2  (-128, -114.3) | -55.6%  (-56.9, -54.1) | -72.9  (-80.3, -65.5) | -34.5%  (-36.7, -32.1) | -17.2  (-25.1, -9.27) | -8.37%  (-11.8, -4.70) | 63.8  (52.5, 75.1) | 18.8%  (15.0, 22.9) |
| Crohn's Disease | -3.37  (-4.37, -2.38) | -17.7%  (-21.8, -13.2) | 0.24  (-1.10, 1.59) | 1.30%  (-5.47, 9.11) | 1.25  (-0.58, 3.08) | 6.72%  (-2.84, 18.4) | 1.91  (-1.54, 5.36) | 6.00%  (-4.34, 18.9) |
| Dementia | -44.3  (-50.5, -38.2) | -22.8%  (-25.2, -20.3) | -9.91  (-16.7, -3.10) | -5.09%  (-8.29, -1.65) | 0.66  (-6.63, 7.95) | 0.34%  (-3.28, 4.24) | 15.9  (5.41, 26.4) | 4.66%  (1.54, 7.98) |
| Depression | -273.6  (-301.6, -245.6) | -29.6%  (-31.7, -27.4) | -174.1  (-202.9, -145.4) | -18.3%  (-20.7, -15.8) | -236.9  (-266.4, -207.4) | -24.3%  (-26.5, -21.9) | -609.3  (-649.7, -568.8) | -34.4%  (-35.9, -32.9) |
| Diabetes Mellitus Type 2 | -100.7  (-128.8, -72.6) | -21.8%  (-26.3, -16.7) | 12.5  (-20.8, 45.9) | 2.71%  (-4.21, 10.7) | 62.0  (22.2, 101.8) | 13.4%  (4.42, 24.1) | 177.4  (116.0, 238.8) | 22.0%  (13.4, 32.0) |
| Epilepsy | -5.13  (-6.87, -3.39) | -11.7%  (-15.1, -8.08) | -2.10  (-3.96, -0.23) | -4.81%  (-8.73, -0.55) | -2.88  (-5.17, -0.59) | -6.60%  (-11.3, -1.42) | -2.93  (-6.35, 0.49) | -3.85%  (-7.99, 0.68) |
| Heart Failure | -48.0  (-54.5, -41.5) | -19.0%  (-21.0, -16.9) | -4.29  (-11.5, 2.96) | -1.65%  (-4.32, 1.17) | 1.29  (-7.45, 10.0) | 0.49%  (-2.73, 3.93) | 4.65  (-8.65, 18.0) | 0.98%  (-1.78, 3.90) |
| Multiple Sclerosis | -0.89  (-1.32, -0.46) | -11.4%  (-16.1, -6.21) | -0.09  (-0.55, 0.36) | -1.22%  (-6.70, 4.95) | -0.46  (-1.03, 0.10) | -5.95%  (-12.4, 1.45) | -0.85  (-1.68, -0.01) | -6.29%  (-11.8, -0.09) |
| Osteoporosis | -63.4  (-68.2, -58.6) | -36.8%  (-38.6, -35.0) | -29.7  (-34.6, -24.9) | -17.1%  (-19.3, -14.7) | -17.5  (-22.4, -12.6) | -9.9%  (-12.3, -7.32) | -27.7  (-34.3, -21.2) | -8.83%  (-10.7, -6.88) |
| Polymyalgia Rheumatica | -3.97  (-6.19, -1.75) | -8.09%  (-12.1, -3.73) | -2.22  (-4.95, 0.52) | -4.65%  (-9.81, 1.15) | -4.03  (-7.22, -0.83) | -8.69%  (-14.6, -1.93) | -0.83  (-5.91, 4.25) | -1.07%  (-7.16, 5.87) |
| Psoriasis | -53.8  (-57.7, -49.8) | -43.8%  (-45.5, -41.9) | -29.9  (-33.9, -25.8) | -24.8%  (-27.3, -22.2) | -11.9  (-16.1, -7.72) | -10.1%  (-13.2, -6.80) | -3.07  (-8.80, 2.67) | -1.54%  (-4.29, 1.38) |
| Rheumatoid Arthritis | -10.0  (-11.9, -8.20) | -24.5%  (-27.7, -21.0) | -4.35  (-6.22, -2.48) | -10.6%  (-14.5, -6.35) | -3.62  (-5.53, -1.71) | -8.85%  (-12.9, -4.38) | -1.88  (-4.49, 0.72) | -2.62%  (-6.03, 1.05) |
| Stroke/TIA | -17.5  (-22.1, -12.9) | -7.06%  (-8.75, -5.31) | 5.92  (1.09, 10.8) | 2.39%  (0.43, 4.43) | 8.95  (2.95, 15.0) | 3.68%  (1.18, 6.30) | 22.1  (13.2, 31.0) | 5.28%  (3.09, 7.56) |
| Ulcerative Colitis | -3.96  (-5.29, -2.62) | -13.4%  (-17.1, -9.26) | 0.11  (-1.28, 1.51) | 0.38%  (-4.14, 5.35) | 1.66  (0.20, 3.11) | 5.60%  (0.66, 11.1) | 4.88  (2.85, 6.91) | 9.41%  (5.29, 13.9) |

Absolute and relative differences between expected and observed incidence rates (IR) for 19 long-term conditions, shown separately for periods after the onset of the COVID-19 pandemic in England. Expected incidence rates were modelled using seasonal autoregressive integrated moving averages (SARIMA), utilising data from April, 2016, to February, 2020. COPD: chronic obstructive pulmonary disease; TIA: transient ischaemic attack.

### **Supplementary Methods:** Autoregressive Integrated Moving Average (ARIMA) modelling

We used Autoregressive Integrated Moving Average (ARIMA) models to analyse time-series of monthly incidence rates for the studied long-term conditions, accounting for autocorrelation and seasonality trends within these data. ARIMA (*p,d,q*) models combine an autoregressive component (where *p* is the number of autoregressive terms), a differencing component (where *d* is the order of differencing), and a moving-average component to induce stationarity (where *q* is the order of the moving-average). For time-series with underlying seasonality, Seasonal ARIMA (SARIMA) can be used to account for these trends. SARIMA models are specified as (*p,d,q*)x(*P,D,Q*)*S*, where *P* and *Q* are the autoregressive and moving-average components of the seasonal model, *D* is the seasonal order of differencing, and *S* is the seasonality in months.

We used the *auto.arima()* function within the *Forecast* package in R to identify SARIMA terms that produced the best-fitting models for each long-term condition, based upon the approaches outlined by Schaffer *et al.* and Qi *et al.^1-3^* Monthly data from April 2016 to February 2020 (i.e. prior to the onset of the COVID-19 pandemic) on age and sex-standardised incidence rates per 100,000 population were fitted using iterative, non-stepwise selection of differencing, seasonality, autoregressive, and moving-average terms to minimise the AIC (Akaike Information Criterion) and BIC (Bayesian Information Criterion). Verification of model selection was performed by visualising plots of residuals against time, residuals against fitted values, normal quantile plots, autocorrelation and partial autocorrelation plots, in addition to performing Ljung-Box tests for white noise and residual autocorrelation. For models where there were clear violations of assumptions of residual normality, constant variance and/or residual autocorrelation, alternative SARIMA terms were explored, and model fit and diagnostics were re-evaluated. SARIMA model specifications for each long-term condition are listed below.

|  | **SARIMA terms**  **(p, d, q) (P, D, Q)[S]** | **AIC** | **BIC** | **Sigma^2^ estimate** | **Log likelihood** |
| --- | --- | --- | --- | --- | --- |
| Asthma | (0,0,0)(0,1,1)[12] | 147.01 | 150.12 | 3.08 | -71.5 |
| Atopic Dermatitis | (0,1,1)(0,1,0)[12] | 77.59 | 80.65 | 0.507 | -36.80 |
| Coronary Heart Disease | (0,0,0)(1,1,0)[12] with drift | 97.99 | 102.66 | 0.763 | -45.99 |
| Chronic Kidney Disease | (0,1,2)(0,1,0)[12] | 177.68 | 182.26 | 9.36 | -85.84 |
| Coeliac Disease | (0,0,0)(0,1,1)[12] | -6.87 | -3.76 | 0.034 | 5.43 |
| COPD | (0,0,0)(0,1,1)[12] with drift | 108.96 | 113.62 | 1.02 | -51.48 |
| Crohn's Disease | (0,1,2)(1,1,0)[12] | -27.05 | -20.95 | 0.018 | 17.53 |
| Dementia | (3,0,0)(0,1,1)[12] | 96.53 | 104.30 | 0.668 | -43.26 |
| Depression | (0,0,0)(0,1,1)[12] with drift | 209.26 | 213.92 | 16.11 | -101.63 |
| Depression (sensitivity) | (0,0,0)(0,1,1)[12] with drift | 226.76 | 231.43 | 28.98 | -110.38 |
| Diabetes Mellitus Type 2 | (0,0,3)(1,1,0)[12] | 180.60 | 188.38 | 7.16 | -85.30 |
| Epilepsy | (0,0,0)(1,1,0)[12] | 12.63 | 15.74 | 0.066 | -4.31 |
| Heart Failure | (0,0,0)(1,1,0)[12] with drift | 103.64 | 108.31 | 0.915 | -48.82 |
| Multiple Sclerosis | (0,0,1)(1,1,0)[12] | -85.02 | -80.35 | 0.0036 | 45.51 |
| Osteoporosis | (0,0,0)(0,1,1)[12] with drift | 86.70 | 91.36 | 0.452 | -40.35 |
| Polymyalgia Rheumatica | (0,1,1)(2,1,0)[12] | 32.40 | 38.51 | 0.103 | -12.20 |
| Psoriasis | (0,0,0)(0,1,1)[12] with drift | 71.98 | 76.65 | 0.321 | -32.99 |
| Rheumatoid Arthritis | (0,0,0)(0,1,1)[12] | 17.56 | 20.67 | 0.068 | -6.78 |
| Stroke | (0,0,0)(1,1,0)[12] with drift | 82.71 | 87.38 | 0.456 | -38.36 |
| Ulcerative Colitis | (0,0,0)(0,1,1)[12] | -4.50 | -1.39 | 0.038 | 4.25 |

Differences between observed incidence rates and expected incidence rates (fitted using SARIMA) were reported for each long-term condition for the following time-periods: full post-pandemic study period (March, 2020, to November, 2024); first year of the pandemic (March, 2020, to February, 2021); second year (March, 2021, to February, 2022); third year (March, 2022, to February, 2023); and from March, 2023, to November, 2024. Differences were reported in absolute and relative terms for each condition, along with 95% prediction intervals. Absolute differences in incident diagnoses with relation to the full population of England were estimated by applying absolute differences in observed vs. expected incidence rates for the study population to mid-year population estimates for England (obtained from the Office for National Statistics).^4^

### **Supplementary Data:** Diagnostic codelists

Incident diagnoses were defined as the first appearance of a diagnostic code for a condition in the primary care or hospitalisation record of individuals from the reference population who did not previously have recorded diagnostic codes for that specific condition.

Primary care diagnoses were defined using the NHS England Primary Care Domain Reference Set codelists, which are collections of SNOMED codes published by NHS England and updated on a regular basis.^5^ These reference sets can be used by clinicians, policy makers, and researchers to collate information on individuals with recorded diagnoses of a particular condition – for example, for use alongside the Quality and Outcomes Framework (QOF) business rules – thereby facilitating comparisons between disparate practices and studies over time.

A comparable set of reference codelists do not exist for hospitalisations, which are based upon the ICD-10 coding system. As such, clinicians within the author team generated a series of ICD-10 codelists for each studied condition, based upon their clinical expertise and with reference to other published codelists (for example, OpenCodelists, and the London School for Hygiene and Tropical Medicine Data Compass). Hospitalisations where relevant diagnostic codes were listed as the primary cause for that admission were included in estimates of incidence and prevalence; however, hospitalisations where diagnostic codes were listed in secondary positions were not included, due to less reliable coding. For example, the inclusion of secondary admission codes for rheumatoid arthritis resulted in incidence rates that were 3-fold higher than estimates obtained from previous population-level studies.^6,7^

Prevalent diagnoses were defined as individuals from the reference population who had prevalent diagnostic codes for a condition in either their primary care or hospitalisation record. For several conditions (defined below), SNOMED codes representing resolved diagnoses of a particular condition also exist – i.e. suggesting the condition is no longer present. Individuals who had resolved codes for a diagnosis that was not superseded by another non-resolved diagnostic code within the study period were not classed as prevalent diagnoses.

**Primary care codelists (SNOMED):**

- https://www.opencodelists.org/codelist/nhsd-primary-care-domain-refsets/ast_cod/3a005293
- https://www.opencodelists.org/codelist/nhsd-primary-care-domain-refsets/astadmsn_cod/3f5c6982
- https://www.opencodelists.org/codelist/nhsd-primary-care-domain-refsets/astres_cod/530fc5c8
- https://www.opencodelists.org/codelist/nhsd-primary-care-domain-refsets/atopic-dermatitis-codes/5816a06b
- https://www.opencodelists.org/codelist/nhsd-primary-care-domain-refsets/chd_cod/0f32f87c
- https://www.opencodelists.org/codelist/nhsd-primary-care-domain-refsets/ckdatrisk2_cod/4529f7b4
- https://www.opencodelists.org/codelist/nhsd-primary-care-domain-refsets/ckdres_cod/305aab93
- https://www.opencodelists.org/codelist/nhsd-primary-care-domain-refsets/coeliac-disease-codes/22e654c2
- https://www.opencodelists.org/codelist/nhsd-primary-care-domain-refsets/copd_cod/3a5ef7dc
- https://www.opencodelists.org/codelist/nhsd-primary-care-domain-refsets/copdadmsn_cod/35d15b44
- https://www.opencodelists.org/codelist/nhsd-primary-care-domain-refsets/copdres_cod/67806ca0
- https://www.opencodelists.org/codelist/nhsd-primary-care-domain-refsets/crohns-disease-codes/3f6f3b7c
- https://www.opencodelists.org/codelist/nhsd-primary-care-domain-refsets/dem_cod/21b792ed
- https://www.opencodelists.org/codelist/nhsd-primary-care-domain-refsets/depr_cod/477e1261
- https://www.opencodelists.org/codelist/nhsd-primary-care-domain-refsets/depres_cod/1369b693
- https://www.opencodelists.org/codelist/nhsd-primary-care-domain-refsets/depsupp_cod/74fe2c6e
- https://www.opencodelists.org/codelist/user/markdrussell/depression_broad/68217c3b/
- https://www.opencodelists.org/codelist/nhsd-primary-care-domain-refsets/dmtype2audit_cod/130b114a
- https://www.opencodelists.org/codelist/nhsd-primary-care-domain-refsets/dmres_cod/62d83746
- https://www.opencodelists.org/codelist/nhsd-primary-care-domain-refsets/epil_cod/28ffb643
- https://www.opencodelists.org/codelist/nhsd-primary-care-domain-refsets/epilres_cod/26b50493
- https://www.opencodelists.org/codelist/nhsd-primary-care-domain-refsets/hf_cod/70b49c5d
- https://www.opencodelists.org/codelist/nhsd-primary-care-domain-refsets/hflvsd_cod/44634425
- https://www.opencodelists.org/codelist/nhsd-primary-care-domain-refsets/hfres_cod/627991fd
- https://www.opencodelists.org/codelist/nhsd-primary-care-domain-refsets/multiple-sclerosis-codes/5ac96061
- https://www.opencodelists.org/codelist/nhsd-primary-care-domain-refsets/osteo_cod/1dd61122
- https://www.opencodelists.org/codelist/user/markdrussell/osteoporosis-resolved/02d12889
- https://www.opencodelists.org/codelist/nhsd-primary-care-domain-refsets/polymyalgia-rheumatica-pmr-codes/5563e075
- https://www.opencodelists.org/codelist/nhsd-primary-care-domain-refsets/adult-and-child-psoriasis-codes/325020f7
- https://www.opencodelists.org/codelist/nhsd-primary-care-domain-refsets/rheumatoid-arthritis-disorders/04d006ea
- https://www.opencodelists.org/codelist/nhsd-primary-care-domain-refsets/strk_cod/78779ff3
- https://www.opencodelists.org/codelist/nhsd-primary-care-domain-refsets/tia_cod/1625df86
- https://www.opencodelists.org/codelist/nhsd-primary-care-domain-refsets/ulcerative-colitis-uc-codes/518bc7a7
- https://www.opencodelists.org/codelist/opensafely/ethnicity-snomed-0removed/22911876

**Secondary care codelists (ICD-10):**

- https://www.opencodelists.org/codelist/user/markdrussell/asthma-secondary-care/2a250f1b
- https://www.opencodelists.org/codelist/user/markdrussell/atopic-dermatitis-secondary-care/645c3567
- https://www.opencodelists.org/codelist/user/markdrussell/coronary-heart-disease-secondary-care/11159be6
- https://www.opencodelists.org/codelist/user/markdrussell/chronic-kidney-disease-secondary-care/167b1bd2
- https://www.opencodelists.org/codelist/user/markdrussell/coeliac-secondary-care/001e6893
- https://www.opencodelists.org/codelist/user/markdrussell/COPD_admission/43348250
- https://www.opencodelists.org/codelist/user/markdrussell/crohns-disease-secondary-care/7ab8e8a6
- https://www.opencodelists.org/codelist/user/markdrussell/dementia-secondary-care/45e74246
- https://www.opencodelists.org/codelist/user/markdrussell/depression-secondary-care/192ddbc8
- https://www.opencodelists.org/codelist/user/markdrussell/type-2-diabetes-secondary-care/2f8a8f07
- https://www.opencodelists.org/codelist/user/markdrussell/epilepsy-secondary-care/13c85bdc
- https://www.opencodelists.org/codelist/user/markdrussell/heart-failure-secondary-care/5ef6b57b
- https://www.opencodelists.org/codelist/user/markdrussell/multiple-sclerosis-secondary-care/2cd7cf11
- https://www.opencodelists.org/codelist/user/markdrussell/osteoporosis-secondary-care/4b4cc232
- https://www.opencodelists.org/codelist/user/markdrussell/polymyalgia-rheumatica-pmr-secondary-care/670ef55d
- https://www.opencodelists.org/codelist/user/markdrussell/psoriasis-secondary-care/7d6ba89c
- https://www.opencodelists.org/codelist/user/markdrussell/rheumatoid-arthritis-secondary-care/245780e9
- https://www.opencodelists.org/codelist/user/markdrussell/stroke-and-tia-secondary-care/780628b0
- https://www.opencodelists.org/codelist/user/markdrussell/ulcerative-colitis-secondary-care/61a97571
